# Inflammatory Bowel Disease and Neurodegenerative Disorders: Integrated Evidence from Mendelian Randomization, Shared Genetic Architecture and Transcriptomics

**DOI:** 10.1101/2022.05.23.22275264

**Authors:** Ruijie Zeng, Jinghua Wang, Rui Jiang, Jie Yang, Chunwen Zheng, Huihuan Wu, Zewei Zhuo, Qi Yang, Jingwei Li, Felix W Leung, Weihong Sha, Hao Chen

## Abstract

**Objective:** Published observational studies have revealed the connection between inflammatory bowel disease (IBD) and neurodegenerative disorders, whereas the causality remains largely unclear. Our study aims to assess the causality and identify the shared genetic architecture between IBD and neurodegenerative disorders.

**Design:** A series of two-sample Mendelian randomization analyses were performed to assess the causality between IBD and neurodegenerative disorders (amyotrophic lateral sclerosis [ALS], Alzheimer’s disease [AD], Parkinson’s disease [PD], and multiple sclerosis [MS]). Shared genetic loci and functional interpretation were further investigated for IBD and ALS. The transcriptomic expressions of shared genes were evaluated in patients with IBD and ALS.

**Results:** Genetic predisposition to IBD is associated with lower odds of ALS (odds ratio [OR] 0.96, 95% confidence interval [CI] 0.94 to 0.99). In contrast, IBD is not genetically associated with an increased risk of AD, PD, or MS. Four shared genetic loci (rs6571361, rs10136727, rs7154847, and rs447853) were derived, and *SCFD1, G2E3, HEATR5A* were further identified as novel risk genes with enriched function related to membrane trafficking. *G2E3* was differentially expressed and significantly correlated with *SCFD1* in patients with IBD or ALS.

**Conclusion:** Our study reveals the casually protective role of IBD on ALS, and does not support the causality of IBD on AD, PD, or MS. Our findings indicate possible shared genetic architecture and pathways between IBD and ALS. The altered expressions of shared risk genes might contribute to the susceptibility to IBD and the protective effects for ALS. These results provide insights into the pathogenesis and therapeutics of IBD and neurodegenerative disorders.

**What is already known on this topic:**

- Emerging evidence has supported the communication between the gastrointestinal tract and central nervous system (the “gut-brain axis”).
- Published epidemiological studies have revealed the association between inflammatory bowel disease (IBD) and neurodegenerative disorders.
- The causality remains largely unclear.

**What this study adds:**

- Genetic liability to IBD is associated with a decreased risk of amyotrophic lateral sclerosis (ALS), whereas the susceptibility to IBD does not lead to Alzheimer’s disease, Parkinson’s disease, or multiple sclerosis.
- Shared genetic loci (rs6571361, rs10136727, rs7154847, and rs447853) and risk genes (*SCFD1, G2E3, HEATR5A*) are identified in IBD and ALS.
- Transcriptomic profiles in patients with IBD or ALS indicate that *G2E3* is differentially expressed and significantly correlated with *SCFD1*.

**How this study might affect research, practice or policy:**

- The findings provide insights into the pathogenesis and therapeutics of IBD and neurodegenerative disorders.
- Lower expression of *G2E3* in IBD might serve as a protective factor to ALS.
- Unsubstantiated concerns among patients with IBD could be alleviated.

## 1. Introduction

Inflammatory bowel diseases (IBD), mainly comprised of Crohn’s diseases (CD) and ulcerative colitis (UC), is a group of chronic and relapsing-remitting disorders of the intestine with an undefined etiology. It is estimated that over 1.5 million and 2.5-3 million individuals are living with IBD in North America and Europe, respectively, and the incidence of IBD in newly industrialized countries is rising rapidly [1]. Numerous complications and extraintestinal conditions are frequently accompanied by IBD, which lead to enormous burdens on the quality of life, economy, and society [2].

Neurodegenerative disorders are debilitating diseases characterized by progressive and selective loss of function or structure of neuronal systems [3]. Neurodegenerative disorders include various intractable diseases such as amyotrophic lateral sclerosis (ALS), Alzheimer’s disease (AD), Parkinson’s disease (PD), and multiple sclerosis with no curative therapy [4]. These disorders share insidious onset and exacerbate irreversibly throughout the disease course, principally occurring in the aging population [5]. As one of the leading causes of disability and mortality worldwide, they also generate immense socioeconomic burdens [6].

Emerging evidence has postulated that IBD is caused by dysregulation of microbiome-immune interactions, in which aberrant microbiome or altered immune responses to the microbiome contribute to intestinal inflammation [7]. The gut-brain axis consists of the bidirectional communications between the intestine and the central nervous system (CNS), and such crosstalk indicates the comorbidities of intestinal inflammation and neurological degeneration [8]. Researchers identified that IBD increases the risk of PD in a Danish nationwide cohort involving 7.5 million individuals during a 37-year follow-up [9]. Another study based on the American cohorts demonstrates that a higher incidence of PD is observed among IBD patients [10]. A study based on the Taiwanese National Health Insurance Research Database indicates that IBD is associated with an increased risk of subsequent development of dementia [11]. The increased prevalence of MS among IBD patients is also verified by a recent meta-analysis [12]. An increasing amount of real-life evidence from epidemiology supports their correlations. In addition, shared mechanisms, for example, autophagy, might be involved in the pathogenesis of IBD and neurodegenerative diseases including ALS [13, 14].

However, the previously reported linkages between IBD and neurodegenerative disorders were mainly observational, while the causality remains largely unexplored. Therefore, it is critical to comprehensively investigate the causal effects of IBD on neurodegenerative disorders using a Mendelian randomization (MR) design (**Figure 1**).

**Figure 1.**
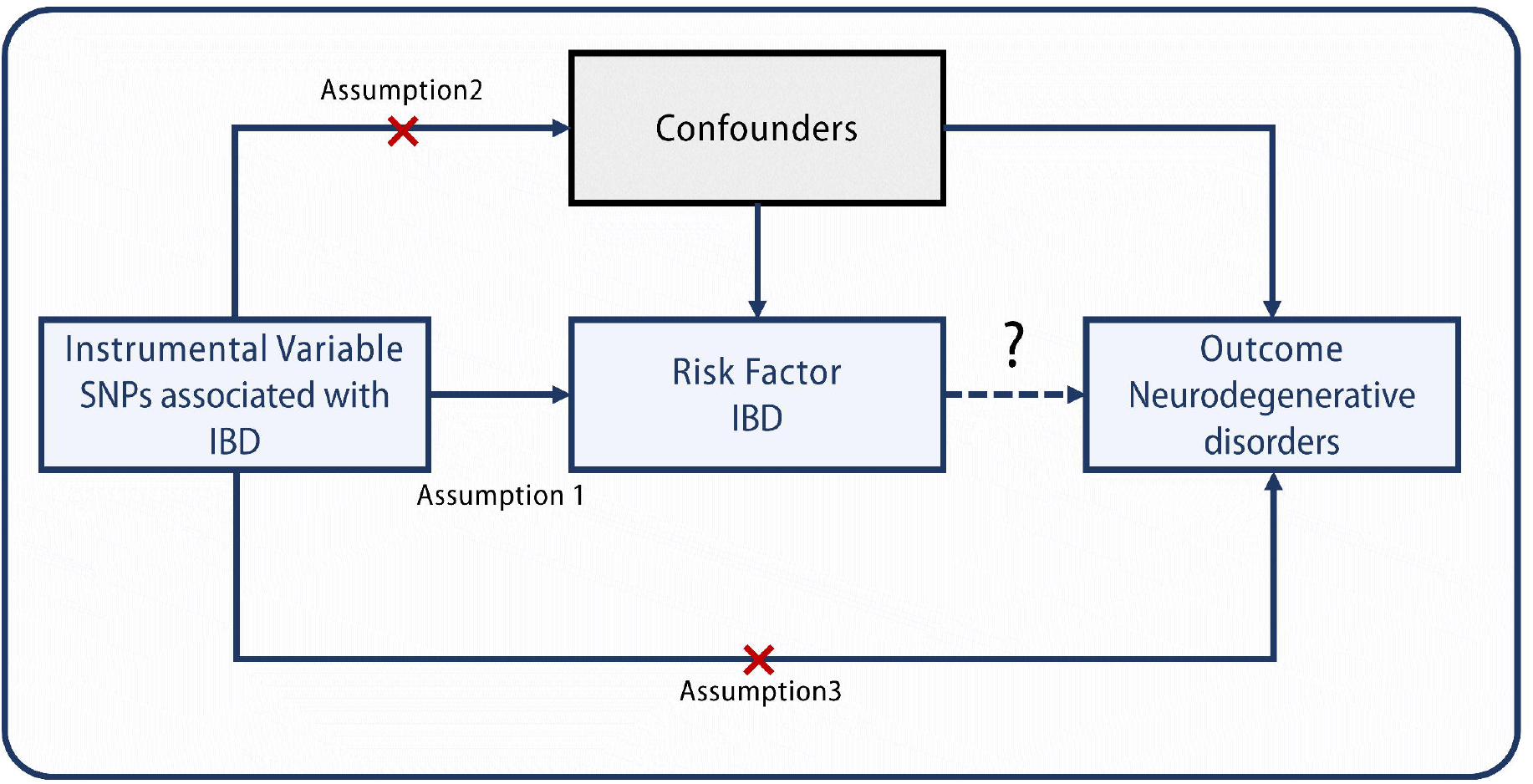
Schematic diagram of the Mendelian randomization (MR) analysis of the causal relationship between inflammatory bowel disease (IBD) and neurodegenerative disorders. The MR is based on three key assumptions: 1) Assumption 1: the instrumental variants (IVs) must be robustly associated with the exposure (IBD); 2) Assumption 2: the association between the IVs and the outcome (neurodegenerative disorders) must be independent of unmeasured confounders; 3) Assumption 3: the IVs influence risk of the outcome (neurodegenerative disorders) only by the exposure, not through other pathways. The causal relationship between IBD and neurodegenerative disorders will not be established if any of the assumptions are infringed. IBD: inflammatory bowel disease; SNP, single-nucleotide polymorphism.

In this study, two-sample MR approach was utilized with large-scale genome-wide association study (GWAS) data to evaluate the potential causal relationship between liability to IBD and neurodegenerative disorders, with a main focus on ALS, AD, PD, and MS. Potential genetic links were explored to further elucidate their correlation, and transcriptomic profiles of the shared risk loci were further evaluated.

## 2. Methods

### 2.1 Data sources

This study relied on publicly available, de-identified and summary-level data mainly from four large-scale cohorts: studies from *Nature Genetics* [15], *Neuron* [16], FinnGen (https://www.finngen.fi/), and the International Parkinson’s Disease Genomics Consortium (IPDGC) [17]. The data for IBD (including UC and CD) from the *Nature Genetics* study were based on 25,042 IBD cases, 12,194 CD cases, 12,366 UC cases, and approximately 35,000 control subjects. The ALS GWAS summary data from the *Neuron* study contained 20,806 cases and 59,804 controls. The data for PD was based on 33,674 cases and 449,056 control subjects. The data for AD and MS were derived from FinnGen, and contained 218,792 individuals (3,899 cases and 214,893 control subjects) and 218,189 individuals (1,048 cases and 217,141 control subjects), respectively. The descriptions of the studies are provided in **Supplementary Table S1**.

### 2.2 Mendelian randomization analysis

Two-sample MR was performed using the TwoSampleMR package [18].The instrumental variables were chosen based on the arbitrary *P* value cut-off. A group of single nucleotide polymorphisms (SNPs) with GWAS significance (*P* < 5 × 10^−8^) associated with each trait were selected. The SNPs were clumped by linkage disequilibrium (LD) with an r^2^ < 0.001 and distance (kb) = 5 000 to ensure that the instruments for the exposure were independent. *F* statistics for each instrument were estimated by *F* = β^2^/SE^2^ [19].

For subsequent analysis, inverse variance weighted (IVW) regression was mainly selected for the inference of causality based on three assumptions: 1) variants are associated with the exposure; 2) variants are independent of confounding factors; 3) variants do not directly affect the outcome [20].

The reverse causality was assessed to evaluate whether neurogenerative disorders were causally associated with IBD.

### 2.3 Sensitivity analysis

For the traits with an IVW *P* < 0.05 and SNP number > 2, heterogeneity tests were performed to evaluate the viability of the effects using Cochran’s Q test and the I^2^ index. Heterogeneity was considered to exist when Cochran’s Q test’s *P* < 0.05 and I^2^ > 50%. MR-Egger and weighted-median (WM) tests were additionally performed to assess the causal effects. MR-Egger regression is based on the assumption that the pleiotropic associates are independent, while it could be inaccurate and largely affected by outlying genetic variants [21]. The WM estimates can provide valid estimates when ≥ 50% of the weight in the analysis comes from the SNVs that are valid instrumental variables [21]. MR test with weighted mode-based estimate (WMBE) was also performed.

Mendelian Randomization Pleiotropy Residual Sum and Outlier (MR-PRESSO) test was performed to identify horizontal pleiotropic outliers [22]. Leave-one-out analysis was used for traits with multiple instruments for assessing whether the causality was driven by one single variant.

The causality was accepted following the criteria as described in the previous publications: IVW was significant and one of the following assumptions was met: 1) no detected heterogeneity with MR-Egger, WM, and WMBE in the same direction; 2) heterogeneity was detected, while it was corrected by MR-PRESSO (<50% of the instruments were considered outliers); 3) heterogeneity existed with MR-PRESSO test detecting >50% of outliers, while MR Egger and WM were significant with the same direction of effect, and WMBE was in the same direction [22, 23].

### 2.4 Genetic correlation analysis and genomic control

Genetic covariance analyzer (GNOVA), which can calculate genetic covariance and estimate genetic correlation according to genetic covariance and heritability, was used to evaluate the genetic correlation between IBD and ALS [24]. The reference data originated from the 1000 Genomes Project European population using default parameters. GNOVA was considered more powerful than conventional cross-trait linkage disequilibrium (LD) score regression (LDSC) and adopted as the method for evaluating genetic correlation in our analysis [24, 25]. ANNOVAR was utilized to annotate SNPs to genic and intragenic regions [26]. PLINK-clump was used to prune the SNPs in LD (r^2^ > 0.2 within 250 kb) [27].

### 2.5 Risk loci identification

Conditional false discovery rate (cFDR) method was used to identify the risk loci showing a strong association with IBD and ALS [28]. cFDRs are characterized as the probability that a certain SNP is falsely positively correlated with the phenotype that the *P* values for both phenotypes (principal and conditional) ≤ the observed *P* values.

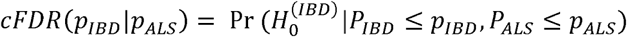

As shown above, p_IBD_ indicates the observed significance that a SNP is associated with IBD, and p_ALS_ indicates the observed strength of association that the same SNP is associated with ALS, while 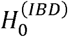 demonstrates the null hypothesis that a SNP is not correlated with IBD. The SNPs with FDR < 0.01 were considered significant. Conjunctional false discovery rate (conjFDR), which was defined as the maximum of the two cFDR statistics, was further calculated to identify loci that were associated with both IBD and ALS [28].

### 2.6 Functional evaluation

The Genotype-Tissue Expression (GTEx) database was used to evaluate the normalized effect size (NES) of the single-tissue cis-expression quantitative trait loci (eQTL) in human tissues [29]. The NES was computed as the effect of the alternative allele compared to the reference allele in human genome reference GRCh38 [29]. The NESs were evaluated in different tissues including adipose, breast, brain, colon, esophagus, heart, lung, muscle, pancreas, skin, spleen, stomach, thyroid, whole blood, artery, nerve, etc. Brain eQTL almanac (Braineac), an online dataset containing data from 10 brain regions obtained from 134 control individuals, was used to investigate eQTL in brain regions, including the cerebellum, frontal cortex, hippocampus, medulla, occipital cortex, putamen, substantia nigra, temporal cortex, thalamus, and white matter [30]. ConsensusPathDB was used to assess the shared pathways associated with IBD and ALS, and 3 ontologic terms (biological process, cellular component, and molecular function) were analyzed [31]. The background gene sets include the shared risk genes identified using the conjunctional FDR and eQTL analyses, and genes located within the human leukocyte antigen (HLA) region were excluded because of the complex LD patterns.

### 2.7 Transcriptomic evaluation of the risk genes

The whole blood expression profiles of ALS patients, IBD patients, and controls were evaluated for the risk genes derived from the above analysis using the datasets GSE112680 and E-MTAB-11349 [32, 33]. The GSE112680 dataset contained transcriptome-wide analysis of whole blood samples derived from 164 ALS cases and 137 control subjects [32]. The E-MTAB-11349 dataset included 323 blood samples from IBD patients and 267 blood samples from control subjects [33](data released on 1 March 2022). The Bioconductor lumi package (v2.44.0) was used for background correction, variance stabilizing transformation, normalization, and quality control of the data [34]. Wilcoxon Rank-Sum test with adjustment by Benjamini-Hochberg method was used to compare the expressions of risk genes between the diseased group and the control group. The ggstatsplot package was used for the evaluation of correlation between gene expression and data visualization [35]. Data were analyzed using R version 4.1.0.

## 3. Results

### 3.1 Instrumental variable selection

After the clumping process, LD-independent SNPs for IBD were derived, and the following conditions were applied to further exclude the listed SNPs: 1) during the extraction of SNPs from the outcomes (ALS, AD, PD, and MS), a certain requested SNP was not identified and a proxy in LD was not able to be found from the outcome; 2) no correction could be performed for ambiguous SNPs or palindromic SNPs with ambiguous strands. Consequently, the SNPs selected as IV for further analysis would be included in those listed in **Supplementary Table S2**. *F* statistics for each IV-exposure association were larger than 10 (ranging from 29.86 to 500.60), and therefore the possibility of weak instrumental variable bias was small in our study.

### 3.2 Two-sample Mendelian randomization analysis for causality of IBD on neurodegenerative disorders

The causal effects of IBD on four major neurodegenerative disorders, including ALS, AD, PD, and MS were explored. **Figure 2** and **Supplementary Figure S1** summarize the estimates of causal effects of IBD on the four neurodegenerative disorders, and **Table 1** demonstrates the detailed information in addition with MR-PRESSO outlier test and the assessment of heterogeneity. Genetically predicted IBD was negatively associated with ALS (IVW [95% CI]: 0.96 [0.94-0.99], *P* = 0.03; all results from the methods were directionally consistent), and no outliers were detected by MR-PRESSO (**Figure 2, Supplementary Figure S1A, Table 1**). The estimates of causal effect were also demonstrated in scatter plots (**Figure 3A**). No evidence of confounding heterogeneity was found by Cochran’s Q test (Q = 113.56, *P* > 0.10; **Table 1**) and leave-one-out test (**Supplementary Figure S2A**). Our results indicated no significant evidence of horizontal pleiotropy for the causality of IBD on ALS (MR-Egger intercept = -0.002, SE = 0.004, *P* = 0.63; MR-PRESSO global test *P* = 0.21; **Supplementary Table S4**). Funnel plots, which provide a visual indication, also demonstrated no horizontal pleiotropy for the causal effect of IBD on ALS (**Supplementary Figure S3A**).

**Figure 2.**
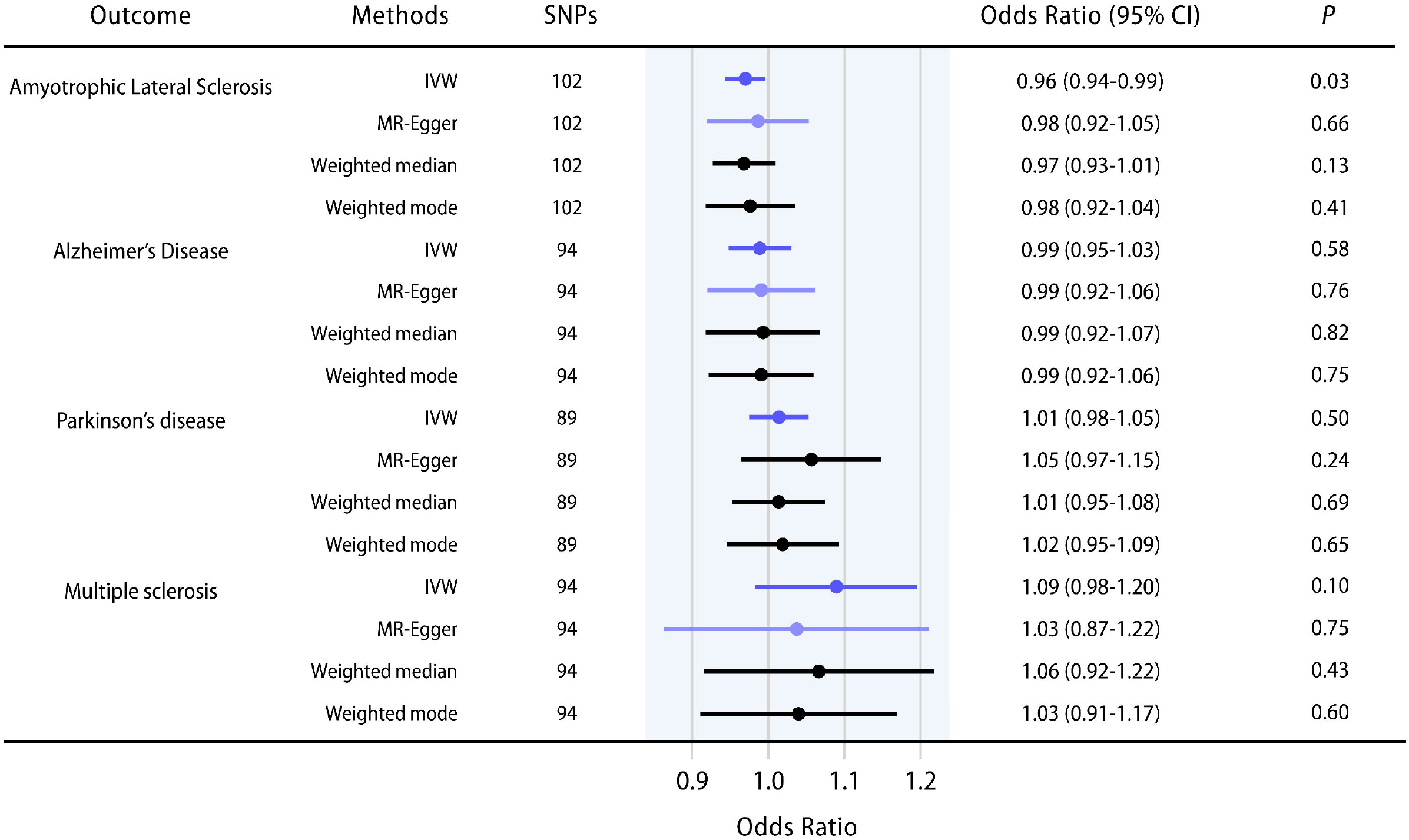
The casual effects of inflammatory bowel disease (IBD) on amyotrophic lateral sclerosis (ALS), Alzheimer’s disease (AD), Parkinson’s disease (PD), and multiple sclerosis (MS). Error bars represent the 95% confidence intervals (CIs) for the estimates. CI: confidence interval; IVW: inverse variance weighting.

**Figure 3.**
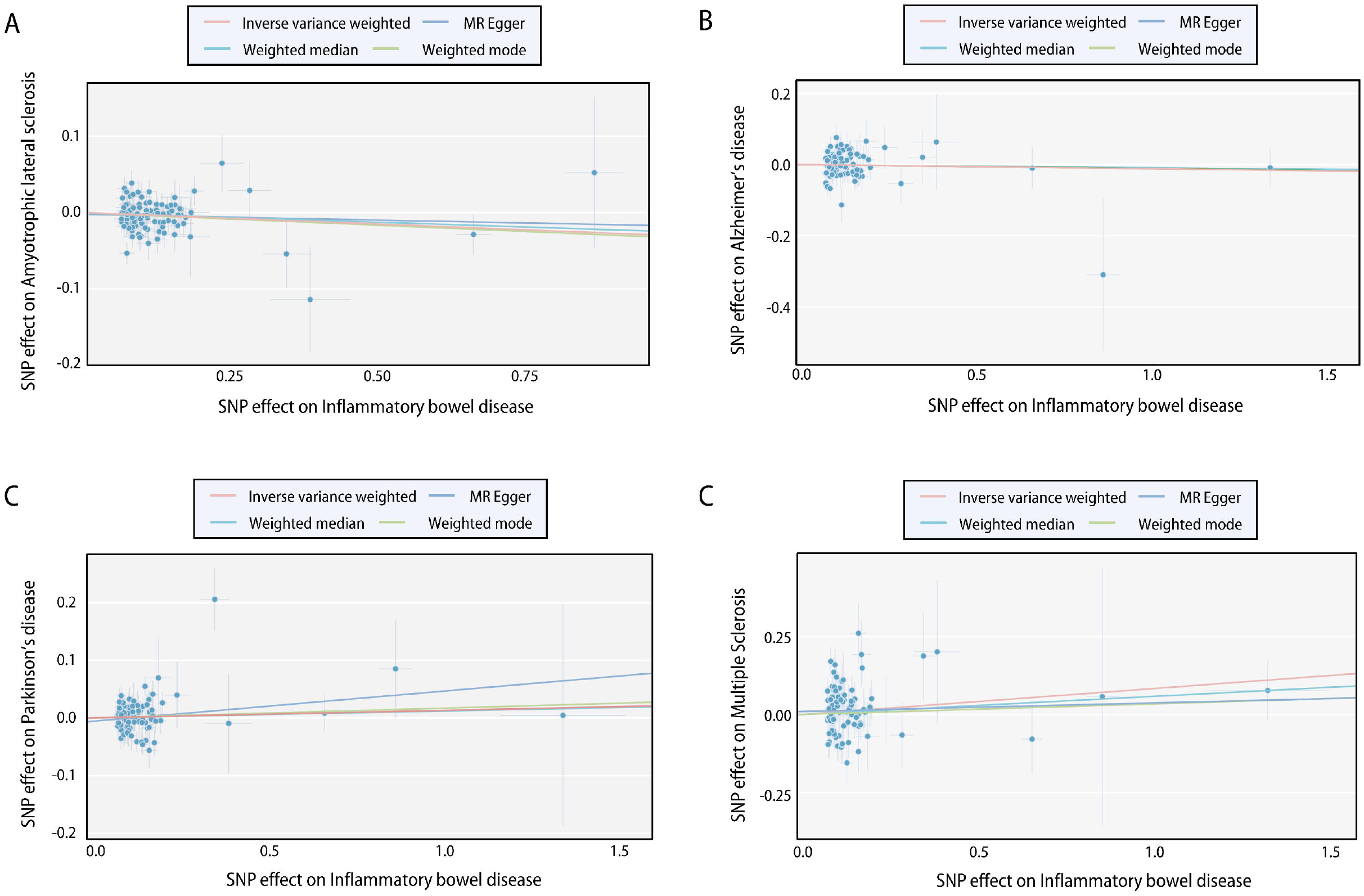
Scatter plots of the causal effect of inflammatory bowel disease (IBD) on neurodegenerative disorders. **A**. IBD on amyotrophic lateral sclerosis (ALS); **B**. IBD on Alzheimer’s disease (AD)**; C**. IBD on Parkinson’s disease (PD); **D**. IBD on multiple sclerosis (MS). The slope of each line indicates the estimation of effects by each method. SNP: single nucleotide polymorphism.

**Table 1.** Mendelian randomization (MR) analysis for the causality of inflammatory bowel diseases (IBD) on neurodegenerative disorders.

| Exposure | Outcome | Method | SNPs | Mendelian randomization |  |  |  | Heterogeneity |  |
| --- | --- | --- | --- | --- | --- | --- | --- | --- | --- |
|  |  |  |  | OR | LL | UL | P | Q | Q_P |
| IBD | ALS | IVW | 102 | 0.97 | 0.94 | 0.99 | 0.03 | 113.56 | 0.19 |
|  |  | MR Egger | 102 | 0.98 | 0.92 | 1.05 | 0.66 | 113.30 | 0.17 |
|  |  | Weighted median | 102 | 0.97 | 0.93 | 1.01 | 0.13 |  |  |
|  |  | Weighted mode | 102 | 0.98 | 0.92 | 1.04 | 0.41 |  |  |
|  |  | MR PERSSO | 102 | 0.97 | 0.94 | 0.99 | 0.03 |  |  |
|  | AD | IVW | 94 | 0.99 | 0.95 | 1.03 | 0.58 | 89.97 | 0.57 |
|  |  | MR Egger | 94 | 0.99 | 0.92 | 1.06 | 0.76 | 89.97 | 0.54 |
|  |  | Weighted median | 94 | 0.99 | 0.92 | 1.07 | 0.82 |  |  |
|  |  | Weighted mode | 94 | 0.99 | 0.92 | 1.06 | 0.75 |  |  |
|  | PD | IVW | 89 | 1.01 | 0.98 | 1.05 | 0.50 | 113.84 | 0.03 |
|  |  | MR Egger | 89 | 1.05 | 0.97 | 1.15 | 0.24 | 112.59 | 0.03 |
|  |  | Weighted median | 89 | 1.01 | 0.95 | 1.08 | 0.69 |  |  |
|  |  | Weighted mode | 89 | 1.02 | 0.95 | 1.09 | 0.65 |  |  |
|  | MS | IVW | 94 | 1.09 | 0.98 | 1.20 | 0.10 | 159.15 | <0.01 |
|  |  | MR Egger | 94 | 1.03 | 0.87 | 1.22 | 0.75 | 158.07 | <0.01 |
|  |  | Weighted median | 94 | 1.06 | 0.92 | 1.22 | 0.43 |  |  |
|  |  | Weighted mode | 94 | 1.03 | 0.91 | 1.17 | 0.60 |  |  |

AD: Alzheimer's disease; ALS: amyotrophic lateral sclerosis; IBD: inflammatory bowel disease; IVW: Inverse variance weighted; LL: lower limits of odds ratio; OR: Odds ratio; PD: Parkinson's disease; MS: multiple sclerosis; SNP: single-nucleotide polymorphism; UL: upper limits of odds ratio.

In contrast, no causality of IBD on AD (IVW [95% CI]: 0.98 [0.94-1.02]; **Figure 2, Supplementary Figure S1B**), PD (IVW [95% CI]: 1.01 [0.98-1.05], *P* = 0.50; **Figure 2, Supplementary Figure S1C**) or MS (IVW [95% CI]: 1.09 [0.98-1.20], *P* = 0.10; **Figure 2, Supplementary Figure S1D**) was found. Scatter plots were shown in **Figure 3B-D**. Forrest plots, leave-one-out test and funnel plots were illustrated in **Supplementary Figure S1B-D, S2B-D and S3B-D**, respectively.

### 3.3 Two-sample Mendelian randomization analysis for causality of CD and UC on neurodegenerative disorders

The two major subtypes of IBD, including CD and UC, were further evaluated for their causal roles on the neurodegenerative disorders. **Supplementary Table S3** summarizes the estimates of causal effects of CD and UC on the four neurodegenerative disorders. Genetically predicted CD was negatively associated with ALS (IVW [95% CI]: 0.97 [0.95-0.99], *P* = 0.02; all results from the methods were directionally consistent), and no outliers were detected by MR-PRESSO (**Supplementary Figure S4A, Supplementary Table S3**). The estimates of causal effect were also demonstrated in scatter plots (**Supplementary S5A**). No evidence of confounding heterogeneity was found by Cochran’s Q test (IVW Q = 77.55, *P* > 0.10; **Supplementary Table S3**) and leave-one-out test (**Supplementary Figure S6A**). Our results indicated no significant evidence of horizontal pleiotropy for the causality of CD on ALS (MR-Egger intercept = -0.008, SE = 0.005, *P* = 0.11; MR-PRESSO global test *P* = 0.46; **Supplementary Table S4**). Funnel plots also indicated no horizontal pleiotropy for the causal effect of CD on ALS (**Supplementary Figure S3D**).

In contrast, no causality of UC on ALS was identified. No causal relationships were found for CD or UC on AD, PD or MS. The risk estimates were demonstrated in **Supplementary Table S3**. Scatter plots were shown in **Supplementary Figure S5B-H**. Forrest plots, leave-one-out tests, and funnel plots were illustrated in **Supplementary Figure S4B-H, S6B-D and S3E-L**, respectively.

### 3.4 Two-sample Mendelian randomization analysis for causality of neurodegenerative disorders on IBD

Significant causal effects of MS on IBD (IVW [95% CI]: 1.11 [1.04-1.18], *P* < 0.01; all results from the methods were directionally consistent) or UC (IVW [95% CI]: 1.18 [1.03-1.34], *P* = 0.02; all results from the methods were directionally consistent; **Supplementary Table S5**) were identified. By contrast, the causality of CD on IBD was not statistically significant (IVW [95% CI]: 1.04 [0.98-1.11], *P* = 0.23; **Supplementary Table S5**). No evidence of causal effects of ALS, AD, and PD on IBD, CD, and UC was identified (**Supplementary Table S5**).

### 3.5 Genetic correlation between IBD and ALS and identification of shared risk loci

Since previously published research has elucidated the causal effect and shared genetic loci of MS on IBD or UC [36], further investigation for shared genetic architecture was mainly focused on MS and ALS in the current study. A significantly negative genetic correlation between IBD and ALS was identified (ρ = -0.047,*P* = 8.60×10^−17^; **Supplementary Table S6**). Conjunctional FDR analysis was further performed to explore genetic variants associated with IBD conditional on ALS. Four risk loci were further identified, namely rs6571361 (*P* = 2.54×10^−7^, FDR = 3.48×10^−2^; *SCFD1*), rs10136727 (*P* = 3.81×10^−7^, FDR = 4.64×10^−2^; *SCFD1*), rs7154847 (*P* = 4.46×10^−7^, FDR = 2.70×10^−2^; *G2E3*), and rs447853 (*P* = 9.58×10^−7^, FDR = 3.45×10^−2^; *G2E3*) (**Table 2**).

**Table 2.**
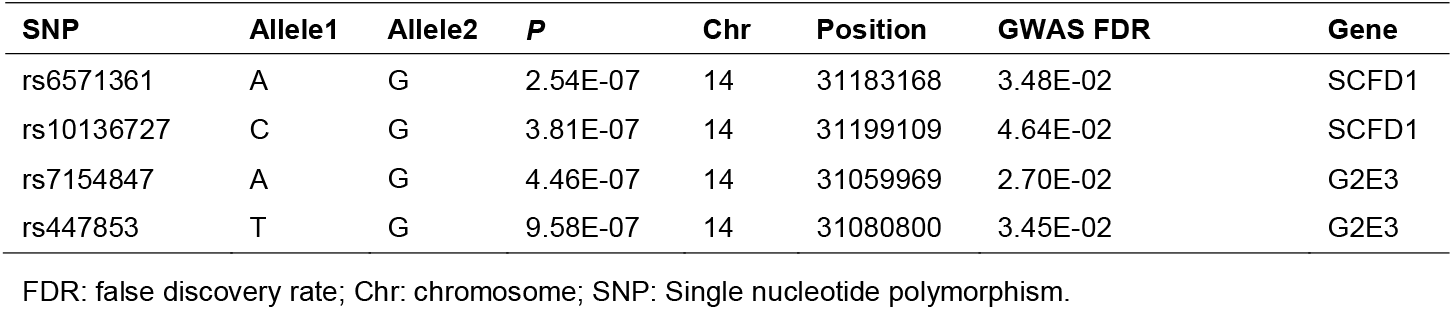
Risk loci associated with inflammatory bowel disease (IBD) conditional on amyotrophic lateral sclerosis (ALS).

### 3.6 Exploration of potential function of shared risk loci

To interpret the function of shared risk loci identified by the conjunction FDR method, cis-eQTL was evaluated in GTEx. The pleiotropic risk loci affect Sec1 Family Domain Containing 1 (SCFD1) and G2/M phase-specific E3 ubiquitin-protein ligase (G2E3) in tissues from both GTEx and Braineac (**Figure 4A, Table 3**). In addition, the pleiotropic risk loci can affect HEAT repeat-containing protein 5A (HEATR5A) in brain tissues (**Table 3**). Based on GTEx analysis, the NES of *SCFD1* was generally higher in brain tissues, whereas that of *SCFD1* was significantly lower in the digestive tract, including esophageal mucosa and colon (**Figure 4A**). The NES of G2E3 was only significantly lower in three tissues, including non-sun-exposed skin, tibial artery, and tibial nerve (**Figure 4A**). Pathway enrichment analysis was further performed to identify biological pathways identified by cis-eQTL analysis. Twenty pathways of biological process were significantly enriched, mainly including vesicle-mediated transport and autophagy-related pathways (*P* < 0.05, **Figure 4B, Supplementary Table S6**). For pathways of cellular components, the Golgi-associated pathways were significantly enriched (*P* < 0.01, **Figure 4B, Supplementary Table S6**). Syntaxin binding pathway was significantly enriched for the pathway of molecular function (*P* < 0.01, **Figure 4B, Supplementary Table S6**). Furthermore, the expressions of *SCFD1, G2E3*, and *HEATR5A* in brain regions were evaluated using the Braineac database. The expression of *SCFD1* was highest in putamen, while that of *SCFD1* was lowest in white matter (Log2 fold change between expression in putamen/white matter = 1.4, *P* = 3.0×10^−12^; **Figure 4C**). *G2E3* expression was highest in putamen, whereas *SCFD1* expression was lowest in substantia nigra (Log2 fold change between expression in putamen/substantia nigra = 1.9, *P* = 5.9 × 10^−24^; **Figure 4C**). The expression of *HEATR5A* was highest in white matter. In contrast, the expression of *HEATR5A* was lowest in cerebellar cortex (Log2 fold change between expression in putamen/white matter = 3.0, *P* = 7.8 × 10^−74^; **Figure 4C**).

**Figure 4.**
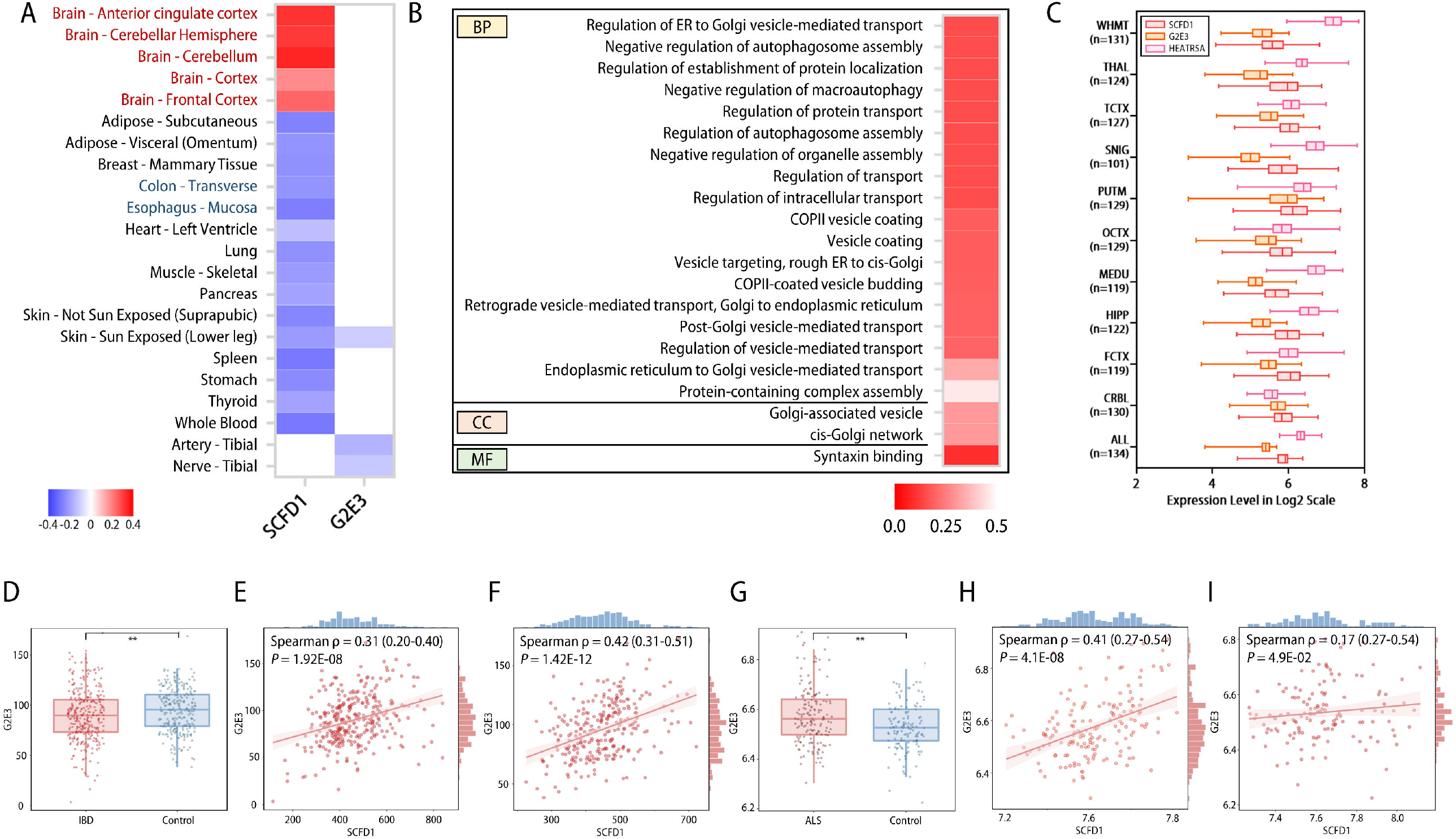
Functional interpretation and transcriptomic analysis of shared risk loci. **A**. Expression quantitative trait loci (eQTL) of shared genetic loci and normalized effect size (NES) of eQTLs in different tissues by GTEx analysis. Blue: negative NES. Red: positive NES. **B**. Enriched pathways related to eQTLs identified by ConsensusPathDB. Deeper color indicates a more significant *P* value. **C**. Expressions of *SCFD1, G2E3*, and *HEATR5A* in different brain regions by Braineac database. **D**. mRNA expressions of *G2E3* in patients with inflammatory bowel disease (IBD) and control subjects. **E-F**. Correlations between *SCFD1* expression and *G2E3* expression in IBD patients **(E)** and control subjects **(F)** from GSE112680. **G**. mRNA expressions of *G2E3* in patients with inflammatory bowel disease (IBD) and control subjects. **H-I**. Correlations between *SCFD1* expression and *G2E3* expression in ALS patients **(H)** and control subjects **(I)** from E-MTAB-11349. ALL, average of all regions; ALS: amyotrophic lateral sclerosis; COPII: coat protein complex II; CRBL, cerebellar cortex; ER: endoplasmic reticulum; FCTX, frontal cortex; HIPP, hippocampus; IBD: inflammatory bowel disease; MEDU, medulla; OCTX, occipital cortex; PUTM, putamen; SNIG, substantia nigra; TCTX, temporal cortex; THAL, thalamus; WHMT, intralobular white matter. * *P* < 0.05, ** *P* < 0.01, *** *P* < 0.001.

**Table 3.** Expression quantitative trait loci (eQTL) indicating the functional effects of shared risk single nucleotide polymorphisms (SNPs) in human brain tissue.

| **Gene Symbol** | **Position** | **Chr** | **start** | **stop** | **P** |
| --- | --- | --- | --- | --- | --- |
| G2E3 | 31199109 | chr14 | 31028362 | 31089250 | **<0.01** |
| G2E3 | 31183168 | chr14 | 31028362 | 31089250 | **0.01** |
| HEATR5A | 31183168 | chr14 | 31760322 | 31889973 | **0.01** |
| SCFD1 | 31183168 | chr14 | 31091515 | 31223811 | **0.02** |
| HEATR5A | 31183168 | chr14 | 31760322 | 31889973 | **0.02** |
| HEATR5A | 31183168 | chr14 | 31760322 | 31889973 | **0.03** |
| HEATR5A | 31183168 | chr14 | 31760322 | 31889973 | **0.04** |
| HEATR5A | 31199109 | chr14 | 31760322 | 31889973 | **0.04** |

3 Chr: chromosome; SNP: Single nucleotide polymorphism.

### 3.7 Differential expression of the risk genes and correlation of gene expressions

The mRNA expressions of *G2E3, SCFD1*, and *HEATR5A* in blood samples of patients with IBD or ALS were further evaluated. The expressions of *G2E3* in blood samples of patients with IBD were significantly lower than that of the control (*P* = 0.0036; **Figure 4D**), and positive correlations between *G2E3* expression and *SCFD1* expression were observed in both patients with IBD (Spearman ρ [95% CI] = 0.31 [0.20-0.40], *P* = 1.92×10^−8^**Figure 4E**) and control subjects (Spearman ρ [95% CI] = 0.42 [0.31-0.51], *P* = 1.42×10^−12^; **Figure 4F**). In contrast, *G2E3* expressions in ALS patients were significantly higher than those of the control group (P = 0.0038; **Figure 4G**). Positive correlations between the expression of *G2E3* and *SCFD1* were identified in both the ALS diseased group (Spearman ρ [95% CI] = 0.41 [0.27-0.54], *P* = 4.1×10^−8^; **Figure 4H**) and the control group (Spearman ρ [95% CI] = 0.41 [0.27-0.54], *P* = 4.9×10^−2^; **Figure 4I**). The expressions of *SCFD1* or *HEATR5A* were not significantly altered, or slightly altered in blood samples of patients with IBD or ALS (**Supplementary Figure S7A-D**). The correlations between expressions of *G2E3* and *HEATR5A, or SCFD1* and *HEATR5A*, were not identified in patients with IBD or ALS, or controls (**Supplementary Figure S7E-L**).

## 4 Discussion

By leveraging large GWAS datasets, our study evaluated the causal effects of IBD on the susceptibility of neurodegenerative disorders using MR design. Our findings provide insights into the causality and shared genetic architecture between IBD and ALS.

To date, the associations between IBD and ALS have not been reported by previous research on epidemiological or genetic aspects. We identified evidence that genetic liability to IBD was significantly associated with a decreased risk of ALS. Moreover, we discovered a significant genetic correlation between IBD and ALS, and plausible mechanisms for the causality between IBD and ALS are discussed as follows.

ALS is a rapidly progressive neuromuscular disease characterized by dysfunction of both upper and lower motor neurons, and most ALS patients die within 3 to 5 years due to respiratory failure [37]. The pathogenesis of ALS is multifactorial and involves complex interactions among diverse environmental and genetic factors. Defects in vesicular trafficking and altered neuronal functions are indicated as one of the pathogenic mechanisms in ALS. Autophagy is a cellular degradation and recycling process mediated by double-membrane vesicles, namely autophagosomes, and can contribute to ALS by leading to neurodegeneration [38]. For IBD, altered morphology of vesicles in colonic mucosa cells increases susceptibility to experimental colitis, and autophagy is regarded as a central issue in IBD development [13, 39]. Based on our findings, the shared risk genes could have novel functional relevance to both IBD and ALS. *SCFD1* plays important roles in mediating vesicle transport and membrane-fusion events, as well as autophagy as indicated by both previous reports and our pathway enrichment results [40]. Recent studies have also identified that *SCFD1* is one of the most significant genes that mediate the risk of ALS [41]. In addition, *G2E3* might function in the regulation of cell cycle and the cellular response to DNA damage [42]. Our findings indicate that the expression of *G2E3* is reduced in patients with IBD, whereas that of *G2E3* is increased in patients with ALS, and the expression of *G2E3* in the two diseases was positively correlated with the expression of *SCFD1*, which could simultaneously exert its functions. Therefore, the decreased expression in patients with IBD might serve as a protective factor for neurodegeneration, contributing to the casual association of IBD with a decreased risk of ALS. Currently, little is known about *G2E3*, and its functions in inflammatory bowel disease or neurodegenerative disorders remain to be explored.

The human gut microbiota is comprised of a community of microorganisms, and a person’s microbiome contains > 150-fold more genes compared to the human genome [8].A wide range of neurotransmitters are produced by gut microbiota and can be traveled across the blood-brain barrier [43]. For example, increased serotonin enhances susceptibility to experimental Crohn’s disease and colitis [44]. In contrast, serotonin receptor dysfunction is involved in the pathogenesis of ALS [45]. In consequence, disrupted intestinal barrier and microbiota in IBD are associated with altered levels of neurotransmitters, while these neurotransmitters could be protective for dysfunctional neurons, which accounts for the decreased risk of ALS.

Intriguingly, our study does not support a causal relationship between IBD and AD or PD, which is contradictory to previous observational studies [9-12]. Since there is a high prevalence of anxiety (32.1%) and depression (25.2%) in patients with IBD, and unsubstantiated concerns would tremendously aggravate their psychological comorbidities, which in turn lead to worse disease outcomes, unnecessary worries should be avoided for patients and clinical practitioners [46]. Although our data indicate that IBD does not lead to MS, the causality of MS on IBD or UC is significant. It is consistent with previously published research, which elucidated that MS has a causal effect on IBD (IVW *P* = 6.62 × 10^−3^) or UC (IVW *P* = 1.65 × 10^−5^) and shared genetic correlations with them [36]. Thus, further investigation for the effects of MS on IBD was not performed in the current study.

The major strength of our study is the two-sample MR design, which limits the confounding and reverse-causality bias in observational studies, with the use of strong instruments. Our results are robust and with no significant evidence of horizontal pleiotropy. In addition, we identified novel SNPs associated with IBD and ALS, which provide plausible explanations for their correlation and deepen current understandings of the disorders. The shared risk genes were validated by transcriptomic analysis. Our study has several limitations. Our study merely involves the European population, which limits the universality of our findings to other ancestries. The causality of UC on ALS is not significant, which might be limited by the scale of the subgroup. Due to the lack of publicly available GWAS summary data or rarity in nature, the causality of IBD on other neurodegenerative disorders was not assessed. Moreover, correction for multiple comparisons might be required. Nevertheless, such correction, for example, the Bonferroni method, is not universally accepted and could artificially create more problems such as an increased likelihood of type II error, leading to the misinterpretation of results [47, 48]. Further investigations with larger and more powerful datasets are warranted.

## 5 Conclusion

Taken together, the current evidence reveals that IBD casually decreases the risk of ALS, whereas the causality of IBD on AD, PD or MS is not supported. Moreover, we identify the possible genetic architecture shared between IBD and ALS, and indicate that modulation of membrane trafficking-related pathways might contribute to their pathogenesis. Decreased expression of *G2E3* in IBD might serve as a protective factor to ALS. Our results provide insights into the causality and shared genetic mechanisms of IBD and ALS, and might contribute to the future development of novel therapeutics.

## Data Availability

GWAS summary statistics are available from the original manuscript of each study in Supplementary Table 1. Code used in this study is available from the corresponding authors upon reasonable request.

## Data sharing statement

GWAS summary statistics are available from the original manuscript of each study in **Supplementary Table 1**. Code used in this study is available from the corresponding authors upon reasonable request.

## Acknowledgement

This work is supported by the National Natural Science Foundation of China (82171698, 82170561, 81300279, 81741067, 82100238), the Natural Science Foundation for Distinguished Young Scholars of Guangdong Province (2021B1515020003), the Guangdong Basic and Applied Basic Research Foundation (2022A1515012081), the Climbing Program of Introduced Talents and High-level Hospital Construction Project of Guangdong Provincial People’s Hospital (DFJH201803, KJ012019099, KJ012021143, KY012021183), and in part by VA Clinical Merit and ASGE clinical research funds (FWL).

## Author contributions

Conceptualization and design: FWL, WHS, HC; Administrative support: FWL, WHS, HC; Funding acquisition: FWL, WHS, HC; Collection and assembly of data: RJZ, JHW, RJ, JY; Data analysis and interpretation: RJZ, JHW, RJ, CWZ, JY, HHW, ZWZ, QY, JWL, KHG; Manuscript writing-original draft: RJZ, JHW, RJ. Manuscript writing-review & editing: CWZ, JY, HHW, ZWZ, QY, JWL, KHG, FWL, WHS, HC. All authors reviewed and approved the final manuscript. All authors had full access to all the data in the study and had final responsibility for the decision to submit for publication.

## References

1 Ng SC, Shi HY, Hamidi N, Underwood FE, Tang W, Benchimol EI, et al. Worldwide incidence and prevalence of inflammatory bowel disease in the 21st century: a systematic review of population-based studies. Lancet 2017;390:2769–78.

2 Peyrin-Biroulet L, Loftus EV, Jr., Colombel JF, Sandborn WJ. Long-term complications, extraintestinal manifestations, and mortality in adult Crohn’s disease in population-based cohorts. Inflamm Bowel Dis 2011;17:471–8.

3 Zhou J, Jangili P, Son S, Ji MS, Won M, Kim JS. Fluorescent Diagnostic Probes in Neurodegenerative Diseases. Adv Mater 2020;32:e2001945.

4 Ong SS, Doraiswamy PM, Lad EM. Controversies and future directions of ocular biomarkers in Alzheimer disease. JAMA neurology 2018;75:650–1.

5 Josephs KA, Ahlskog JE, Parisi JE, Boeve BF, Crum BA, Giannini C, et al. Rapidly progressive neurodegenerative dementias. Arch Neurol 2009;66:201–7.

6 Feigin VL, Nichols E, Alam T, Bannick MS, Beghi E, Blake N, et al. Global, regional, and national burden of neurological disorders, 1990–2016: a systematic analysis for the Global Burden of Disease Study 2016. The Lancet Neurology 2019;18:459–80.

7 Glassner KL, Abraham BP, Quigley EMM. The microbiome and inflammatory bowel disease. J Allergy Clin Immunol 2020;145:16–27.

8 Long-Smith C, O’Riordan KJ, Clarke G, Stanton C, Dinan TG, Cryan JF. Microbiota-Gut-Brain Axis: New Therapeutic Opportunities. Annu Rev Pharmacol Toxicol 2020;60:477–502.

9 Villumsen M, Aznar S, Pakkenberg B, Jess T, Brudek T. Inflammatory bowel disease increases the risk of Parkinson’s disease: a Danish nationwide cohort study 1977-2014. Gut 2019;68:18–24.

10 Peter I, Dubinsky M, Bressman S, Park A, Lu C, Chen N, et al. Anti-Tumor Necrosis Factor Therapy and Incidence of Parkinson Disease Among Patients With Inflammatory Bowel Disease. JAMA Neurol 2018;75:939–46.

11 Zhang B, Wang HE, Bai YM, Tsai SJ, Su TP, Chen TJ, et al. Inflammatory bowel disease is associated with higher dementia risk: a nationwide longitudinal study. Gut 2021;70:85–91.

12 Wang X, Wan J, Wang M, Zhang Y, Wu K, Yang F. Multiple sclerosis and inflammatory bowel disease: A systematic review and meta-analysis. Annals of Clinical and Translational Neurology 2022;9:132–40.

13 Wang S-L, Shao B-Z, Zhao S-B, Chang X, Wang P, Miao C-Y, et al. Intestinal autophagy links psychosocial stress with gut microbiota to promote inflammatory bowel disease. Cell death & disease 2019;10:1–17.

14 Nixon RA. The role of autophagy in neurodegenerative disease. Nature medicine 2013;19:983–97.

15 de Lange KM, Moutsianas L, Lee JC, Lamb CA, Luo Y, Kennedy NA, et al. Genome-wide association study implicates immune activation of multiple integrin genes in inflammatory bowel disease. Nat Genet 2017;49:256–61.

16 Nicolas A, Kenna KP, Renton AE, Ticozzi N, Faghri F, Chia R, et al. Genome-wide Analyses Identify KIF5A as a Novel ALS Gene. Neuron 2018;97:1268–83.e6.

17 Nalls MA, Blauwendraat C, Vallerga CL, Heilbron K, Bandres-Ciga S, Chang D, et al. Identification of novel risk loci, causal insights, and heritable risk for Parkinson’s disease: a meta-analysis of genome-wide association studies. Lancet Neurol 2019;18:1091–102.

18 Hemani G, Zheng J, Elsworth B, Wade KH, Haberland V, Baird D, et al. The MR-Base platform supports systematic causal inference across the human phenome. Elife 2018;7.

19 Li B, Martin EB. An approximation to the F distribution using the chi-square distribution. Computational statistics & data analysis 2002;40:21–6.

20 Burgess S, Butterworth A, Thompson SG. Mendelian randomization analysis with multiple genetic variants using summarized data. Genet Epidemiol 2013;37:658–65.

21 Bowden J, Del Greco MF, Minelli C, Davey Smith G, Sheehan NA, Thompson JR. Assessing the suitability of summary data for two-sample Mendelian randomization analyses using MR-Egger regression: the role of the I2 statistic. Int J Epidemiol 2016;45:1961–74.

22 Verbanck M, Chen CY, Neale B, Do R. Detection of widespread horizontal pleiotropy in causal relationships inferred from Mendelian randomization between complex traits and diseases. Nat Genet 2018;50:693–8.

23 Marini S, Merino J, Montgomery BE, Malik R, Sudlow CL, Dichgans M, et al. Mendelian randomization study of obesity and cerebrovascular disease. Annals of neurology 2020;87:516–24.

24 Lu Q, Li B, Ou D, Erlendsdottir M, Powles RL, Jiang T, et al. A Powerful Approach to Estimating Annotation-Stratified Genetic Covariance via GWAS Summary Statistics. Am J Hum Genet 2017;101:939–64.

25 Bulik-Sullivan BK, Loh PR, Finucane HK, Ripke S, Yang J, Patterson N, et al. LD Score regression distinguishes confounding from polygenicity in genome-wide association studies. Nat Genet 2015;47:291–5.

26 Wang K, Li M, Hakonarson H. ANNOVAR: functional annotation of genetic variants from high-throughput sequencing data. Nucleic Acids Res 2010;38:e164.

27 Purcell S, Neale B, Todd-Brown K, Thomas L, Ferreira MA, Bender D, et al. PLINK: a tool set for whole-genome association and population-based linkage analyses. Am J Hum Genet 2007;81:559–75.

28 Andreassen OA, Djurovic S, Thompson WK, Schork AJ, Kendler KS, O’Donovan MC, et al. Improved detection of common variants associated with schizophrenia by leveraging pleiotropy with cardiovascular-disease risk factors. Am J Hum Genet 2013;92:197–209.

29 Lonsdale J, Thomas J, Salvatore M, Phillips R, Lo E, Shad S, et al. The genotype-tissue expression (GTEx) project. Nature genetics 2013;45:580–5.

30 Ramasamy A, Trabzuni D, Guelfi S, Varghese V, Smith C, Walker R, et al. Genetic variability in the regulation of gene expression in ten regions of the human brain. Nature neuroscience 2014;17:1418–28.

31 Kamburov A, Wierling C, Lehrach H, Herwig R. ConsensusPathDB--a database for integrating human functional interaction networks. Nucleic Acids Res 2009;37:D623–8.

32 Swindell WR, Kruse CP, List EO, Berryman DE, Kopchick JJ. ALS blood expression profiling identifies new biomarkers, patient subgroups, and evidence for neutrophilia and hypoxia. Journal of translational medicine 2019;17:1–33.

33 Brazma A, Parkinson H, Sarkans U, Shojatalab M, Vilo J, Abeygunawardena N, et al. ArrayExpress—a public repository for microarray gene expression data at the EBI. Nucleic acids research 2003;31:68–71.

34 Du P, Kibbe WA, Lin SM. lumi: a pipeline for processing Illumina microarray. Bioinformatics 2008;24:1547–8.

35 Patil I. Visualizations with statistical details: The’ggstatsplot’approach. Journal of Open Source Software 2021;6:3167.

36 Yang Y, Musco H, Simpson-Yap S, Zhu Z, Wang Y, Lin X, et al. Investigating the shared genetic architecture between multiple sclerosis and inflammatory bowel diseases. Nature communications 2021;12:1–12.

37 Chia R, Chiò A, Traynor BJ. Novel genes associated with amyotrophic lateral sclerosis: diagnostic and clinical implications. Lancet Neurol 2018;17:94–102.

38 Chua JP, De Calbiac H, Kabashi E, Barmada SJ. Autophagy and ALS: Mechanistic insights and therapeutic implications. Autophagy 2022;18:254–82.

39 Hino K, Saito A, Asada R, Kanemoto S, Imaizumi K. Increased susceptibility to dextran sulfate sodium-induced colitis in the endoplasmic reticulum stress transducer OASIS deficient mice. PloS one 2014;9:e88048.

40 Huang H, Ouyang Q, Zhu M, Yu H, Mei K, Liu R. mTOR-mediated phosphorylation of VAMP8 and SCFD1 regulates autophagosome maturation. Nature communications 2021;12:1–15.

41 Saez-Atienzar S, Bandres-Ciga S, Langston RG, Kim JJ, Choi SW, Reynolds RH, et al. Genetic analysis of amyotrophic lateral sclerosis identifies contributing pathways and cell types. Science advances 2021;7:eabd9036.

42 Brooks WS, Banerjee S, Crawford DF. G2E3 is a nucleo-cytoplasmic shuttling protein with DNA damage responsive localization. Exp Cell Res 2007;313:665–76.

43 Strandwitz P. Neurotransmitter modulation by the gut microbiota. Brain research 2018;1693:128–33.

44 Haq S, Wang H, Grondin J, Banskota S, Marshall JK, Khan, II, et al. Disruption of autophagy by increased 5-HT alters gut microbiota and enhances susceptibility to experimental colitis and Crohn’s disease. Sci Adv 2021;7:eabi6442.

45 El Oussini H, Scekic-Zahirovic J, Vercruysse P, Marques C, Dirrig-Grosch S, Dieterlé S, et al. Degeneration of serotonin neurons triggers spasticity in amyotrophic lateral sclerosis. Annals of neurology 2017;82:444–56.

46 Barberio B, Zamani M, Black CJ, Savarino EV, Ford AC. Prevalence of symptoms of anxiety and depression in patients with inflammatory bowel disease: a systematic review and meta-analysis. The Lancet Gastroenterology & Hepatology 2021;6:359–70.

47 Perneger TV. What’s wrong with Bonferroni adjustments. Bmj 1998;316:1236–8.

48 Perneger TV. Adjusting for multiple testing in studies is less important than other concerns. Bmj 1999;318:1288.

