## Supplementary material for "Inflammatory Bowel Disease and Neurodegenerative Disorders: Integrated Evidence from Mendelian Randomization, Shared Genetic Architecture and Transcriptomics": Supplementary Materials.pdf

**Supplementary Information**

**Supplementary Figure S1. Forest plots illustrate the effect of inflammatory bowel disease (IBD) on neurodegenerative disorders. A**. IBD on amyotrophic lateral sclerosis (ALS); **B**. IBD on Alzheimer’s disease (AD); **C**. IBD on Parkinson’s disease (PD); **D.** IBD on multiple sclerosis (MS).

**Supplementary Figure S2. Leave-one-out sensitivity analysis for inflammatory bowel disease (IBD) on neurodegenerative disorders.** A. IBD on amyotrophic lateral sclerosis (ALS); B. IBD on Alzheimer’s disease (AD); C. IBD on Parkinson’s disease (PD); **D.** IBD on multiple sclerosis (MS).

**Supplementary Figure S3. Funnel plots evaluating the horizontal heterogeneity for the effect of inflammatory bowel disease (IBD) on neurodegenerative disorders. A.** IBD on amyotrophic lateral sclerosis (ALS); **B.** IBD on Alzheimer’s disease (AD); **C.** IBD on Parkinson’s disease (PD); **D.** IBD on multiple sclerosis (MS); **E.** Crohn’s disease (CD) on ALS; **F**. CD on AD; **G.** CD on PD; **H.** CD on MS; **I**. ulcerative colitis (UC) on ALS; **J.** UC on AD; **K** UC on PD; **L.** UC on MS.

**Supplementary Figure S4. Forest plots demonstrate the effect of Crohn’s disease (CD) and ulcerative colitis (UC) on neurodegenerative disorders. A.** CD on amyotrophic lateral sclerosis (ALS)**; B.** CD on Alzheimer’s disease (AD); **C.** CD on Parkinson’s disease (PD); **D.** CD on multiple sclerosis (MS); **E.** UC on ALS; **F.** UC on AD; **G.** UC on PD; **H.** UC on MS.

**Supplementary Figure S5. Scatter plots of the causal effect of Crohn’s disease (CD) and ulcerative colitis (UC) on neurodegenerative disorders. A.** CD on amyotrophic lateral sclerosis (ALS)**; B.** CD on Alzheimer’s disease (AD); **C.** CD on Parkinson’s disease (PD); **D.** CD on multiple sclerosis (MS); **E.** UC on ALS; **F.** UC on AD; **G.** UC on PD; **H.** UC on MS.

**Supplementary Figure S6. Leave-one-out sensitivity analysis for Crohn’s disease (CD) and ulcerative colitis (UC) on neurodegenerative disorders**. **A.** CD on amyotrophic lateral sclerosis (ALS)**; B.** CD on Alzheimer’s disease (AD); **C.** CD on Parkinson’s disease (PD); **D.** CD on multiple sclerosis (MS); **E.** UC on ALS; **F.** UC on AD; **G.** UC on PD; **H.** UC on MS.

**Supplementary Figure S7. Transcriptomic analysis of shared risk loci. A-B.** mRNA expressions of SCFD1 **(A)** and HEATR5A **(B)** in IBD patients and control subjects from GSE112680. **C-D.** mRNA expressions of SCFD1 **(C)** and HEATR5A **(D)** in ALS patients and control subjects from E-MTAB-11349. **E-F.** Correlations between *G2E3* expression and *HEATR5A* expression in IBD patients **(E)** and control subjects **(F)** from GSE112680. **G-H.** Correlations between *SCFD1* expression and *HEATR5A* expression in IBD patients **(G)** and control subjects **(H)** from GSE112680. **I-J.** Correlations between *G2E3* expression and *HEATR5A* expression in ALS patients **(I)** and control subjects **(J)** from E-MTAB-11349. **K-L.** Correlations between *SCFD1* expression and *HEATR5A* expression in ALS patients **(K)** and control subjects **(L)** from E-MTAB-11349. * $P < 0.05$, ** $P < 0.01$, *** $P < 0.001$, NS: non-significant.

**Supplementary Table S1. Characteristics of data sources used in this study**

**Supplementary Table S2. Detailed information for SNPs in the Mendelian Randomization analysis of inflammatory bowel disease (IBD) on neurodegenerative disorders.**

**Supplementary Table S3. Mendelian randomization (MR) analysis for the causality ulcerative colitis (UC) and Crohn's disease (CD) on neurodegenerative disorders.**

**Supplementary Table S4. Assessment of pleiotropy.**

**Supplementary Table S5. Mendelian randomization (MR) analysis for the causality of neurodegenerative disorders on inflammatory bowel disease (IBD), including ulcerative colitis (UC) and Crohn's disease (CD).**

**Supplementary Table S6. Genetic correlation between inflammatory bowel disease (IBD) and amyotrophic lateral sclerosis (ALS).**

**Supplementary Table S7. Functional enrichment of shared risk loci.**

### Supplementary Figure S1

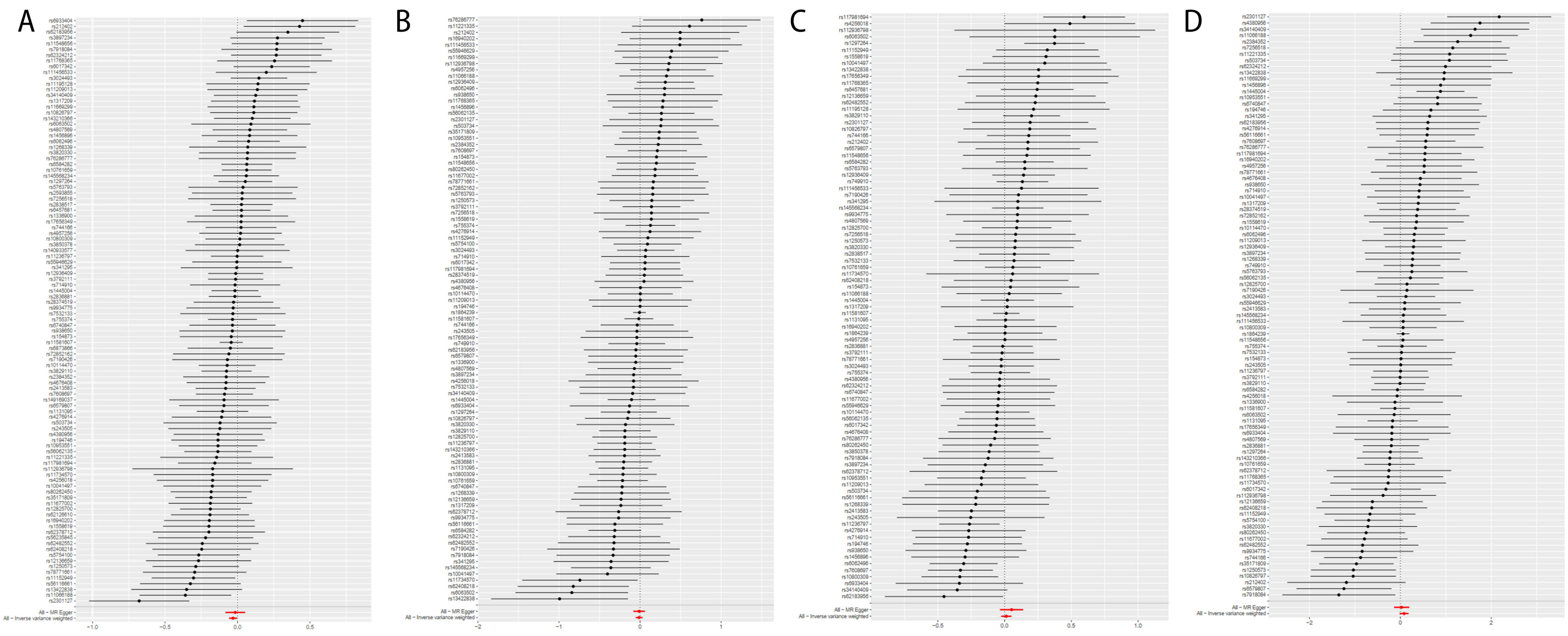

### Supplementary Figure S2

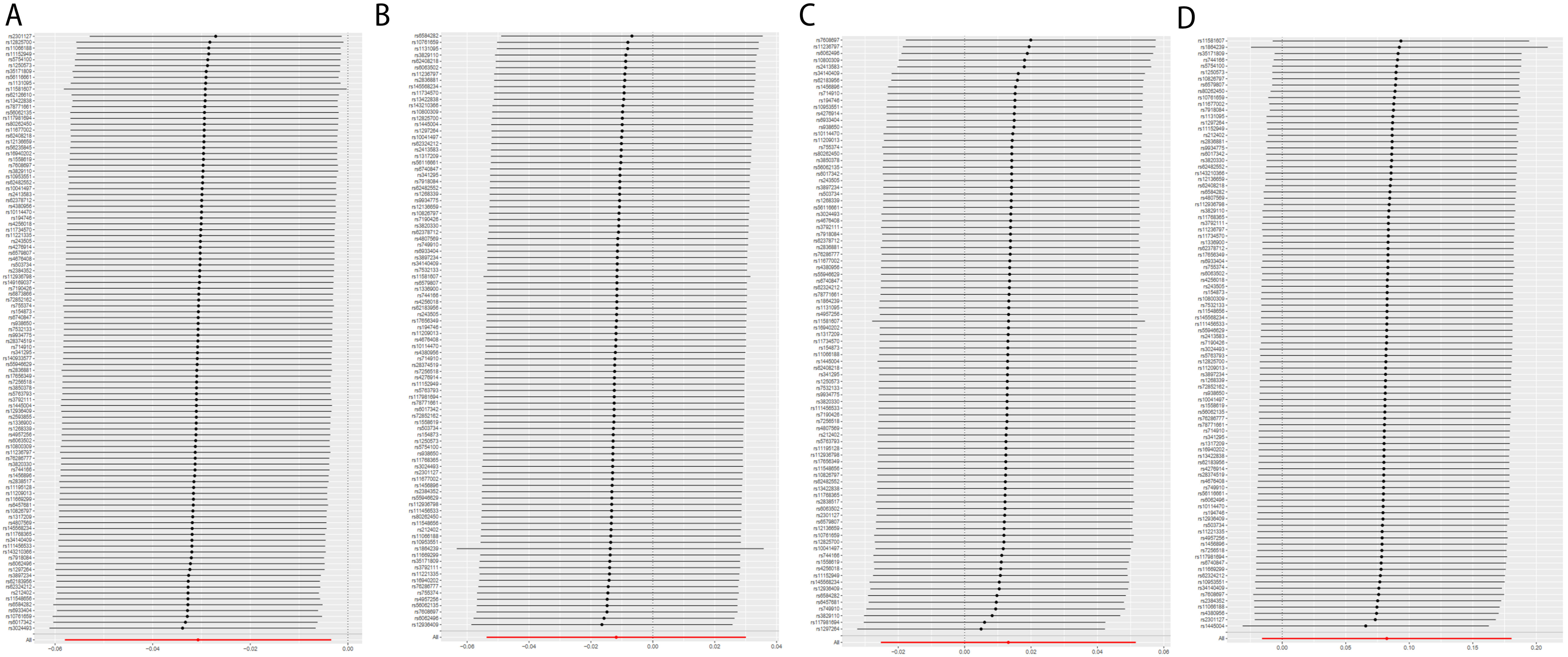

### Supplementary Figure S3

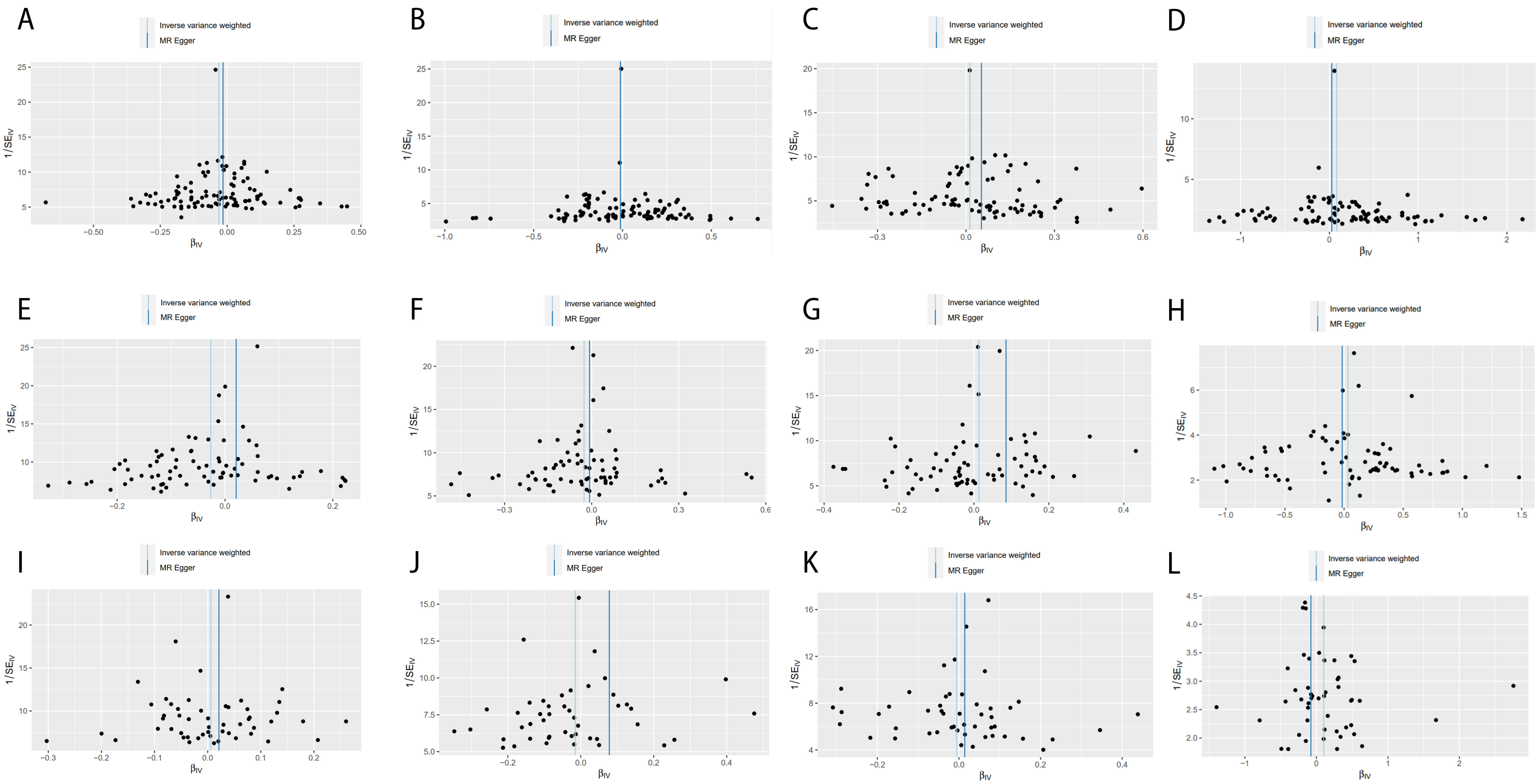

### Supplementary Figure S4

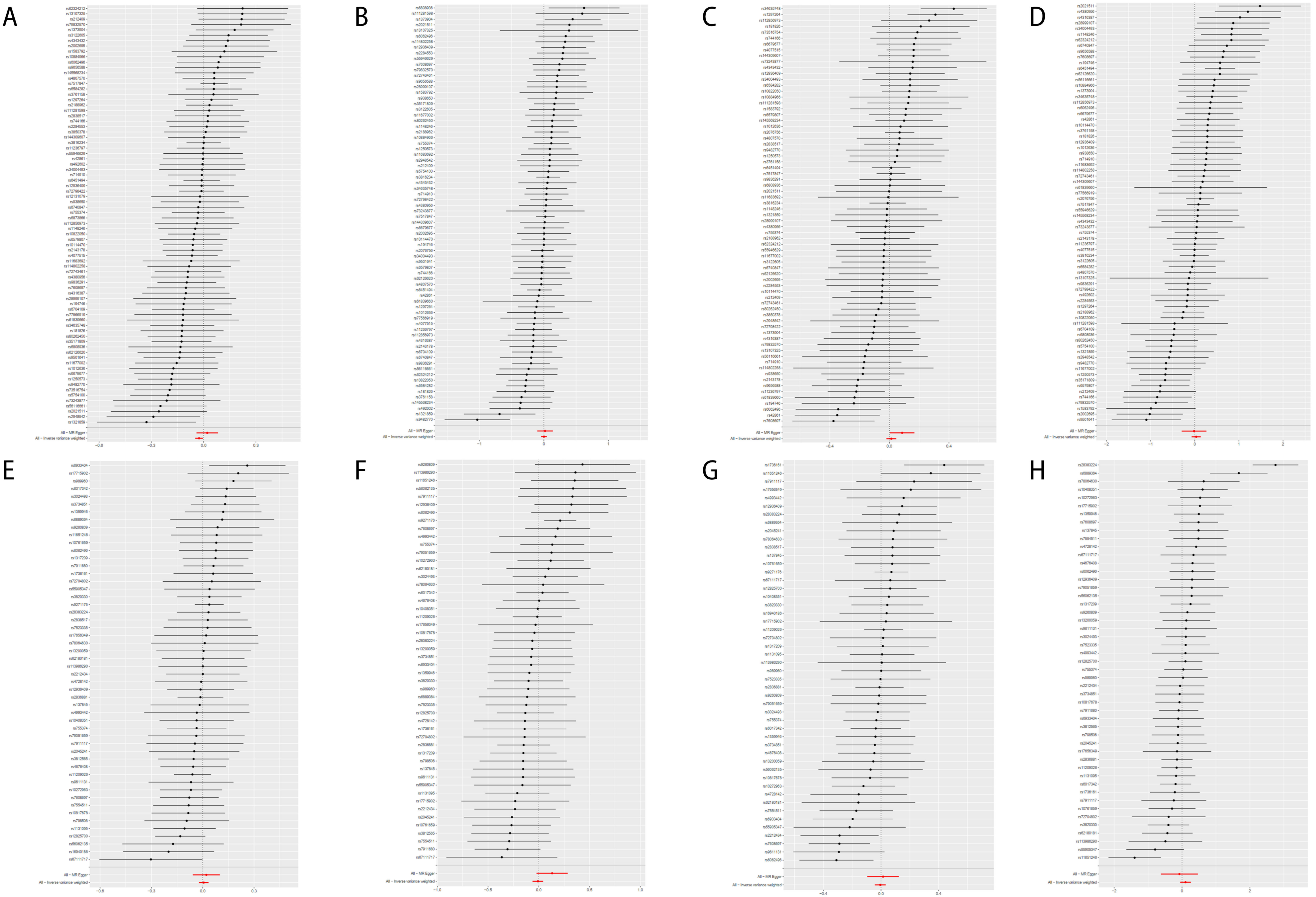

### Supplementary Figure S5

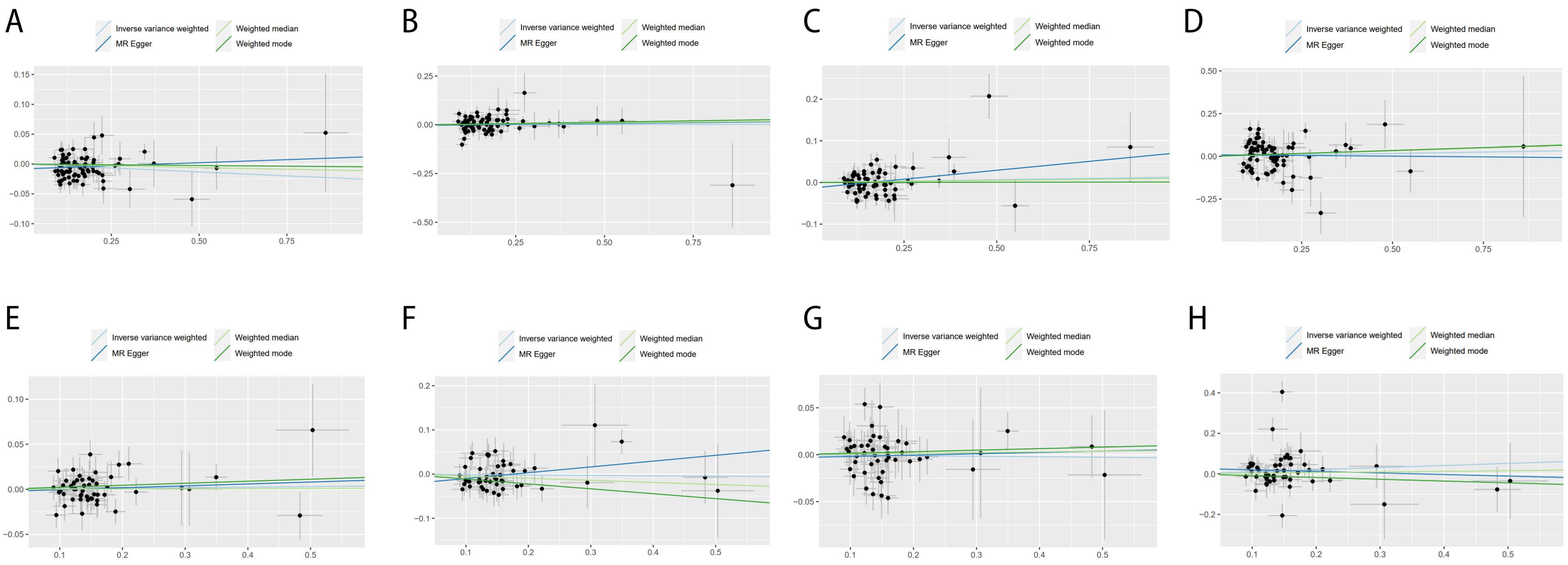

### Supplementary Figure S6

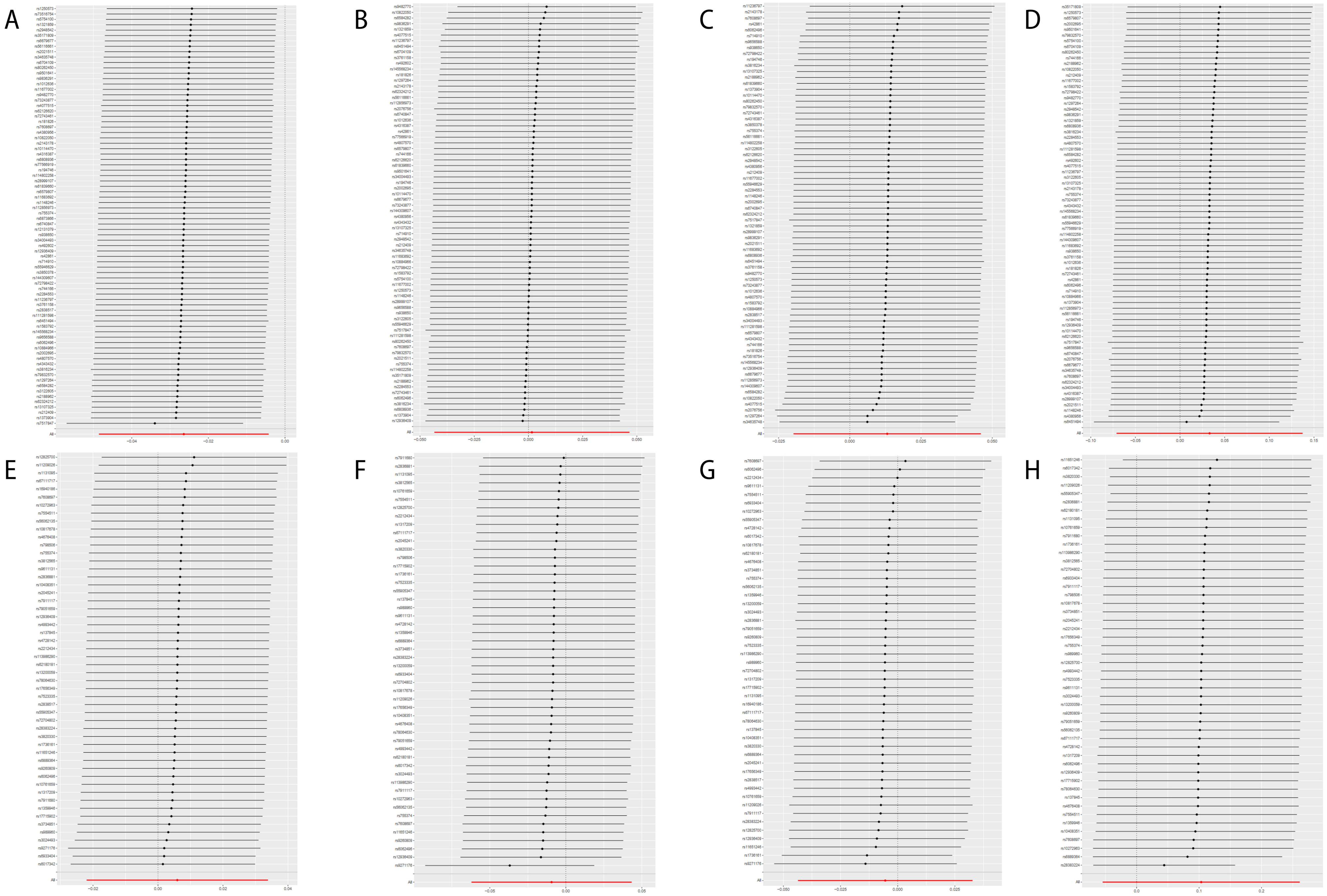

### Supplementary Figure S7

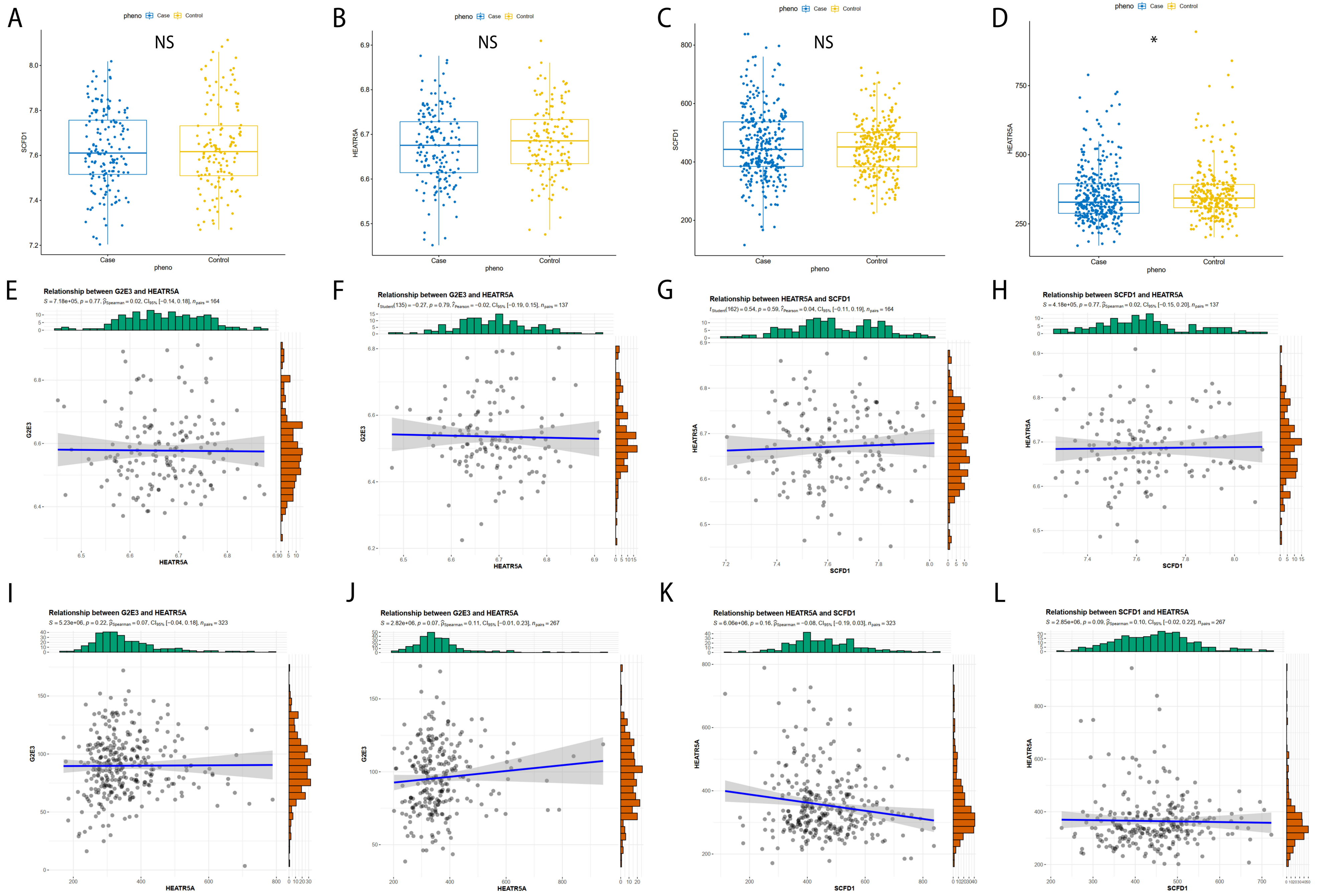

**Supplementary Table S1. Characteristics of data sources used in this study**

| Traits | Data sources | Sample size (cases/controls) | Ancestry | Reference |
|---|---|---|---|---|
| IBD | *Nature Genetics* | 25,042/34,915 | European | de Lange et al., 2017 |
| CD | | 12,194/28,072 | | |
| UC | | 12,366/33,609 | | |
| ALS | *Neuron* | 20,806/59,804 | European | Nicolas et al., 2018 |
| AD | FinnGen | 3,899/214,893 | European | https://www.finngen.fi/ |
| PD | International Parkinson's Disease Genomics Consortium | 33,674/449,056 | European | Nalls et al., 2019 |
| MS | FinnGen | 1,048/217,141 | European | https://www.finngen.fi/ |

AD: Alzheimer’s disease; ALS: amyotrophic lateral sclerosis; CD: Crohn’s disease; IBD: inflammatory bowel disease; MS: multiple sclerosis; PD: Parkinson’s disease; UC: ulcerative colitis.

**Supplementary Table S2. Detailed information for SNPs in the Mendelian Randomization analysis of inflammatory bowel disease (IBD) on neurodegenerative disorders.**

| SNP | Chr | Location | Association with ALS | | | Association with AD | | | Association with PD | | | Association with MS | | |
|---|---|---|---|---|---|---|---|---|---|---|---|---|---|---|
| | | | β | SE | *P* | β | SE | *P* | β | SE | *P* | β | SE | *P* |
| rs10041497 | 5 | 142135017 | -0.17 | 0.17 | 0.31 | -0.40 | 0.32 | 0.21 | 0.30 | 0.24 | 0.20 | 0.40 | 0.58 | 0.49 |
| rs10114470 | 9 | 114785492 | -0.07 | 0.10 | 0.48 | <0.01 | 0.20 | 0.99 | -0.05 | 0.12 | 0.66 | 0.33 | 0.36 | 0.36 |
| rs10761659 | 10 | 62685804 | 0.06 | 0.09 | 0.46 | -0.22 | 0.16 | 0.17 | 0.06 | 0.11 | 0.56 | -0.24 | 0.28 | 0.40 |
| rs10800309 | 1 | 161502368 | 0.02 | 0.12 | 0.89 | -0.21 | 0.21 | 0.32 | -0.33 | 0.15 | 0.02 | 0.06 | 0.38 | 0.88 |
| rs10826797 | 10 | 30401447 | 0.11 | 0.15 | 0.47 | -0.15 | 0.27 | 0.57 | 0.19 | 0.25 | 0.46 | -1.04 | 0.48 | 0.03 |
| rs10953551 | 7 | 107840456 | -0.13 | 0.14 | 0.33 | 0.23 | 0.25 | 0.35 | -0.17 | 0.17 | 0.31 | 0.82 | 0.45 | 0.07 |
| rs11066188 | 12 | 112172910 | -0.36 | 0.16 | 0.03 | 0.33 | 0.30 | 0.27 | 0.03 | 0.20 | 0.86 | 1.55 | 0.53 | <0.01 |
| rs111456533 | 10 | 124750812 | 0.20 | 0.18 | 0.26 | 0.49 | 0.39 | 0.21 | 0.13 | 0.29 | 0.67 | 0.06 | 0.68 | 0.93 |
| rs11152949 | 6 | 106001210 | -0.30 | 0.15 | 0.04 | 0.10 | 0.29 | 0.74 | 0.32 | 0.20 | 0.10 | -0.67 | 0.51 | 0.19 |
| rs11195128 | 10 | 110426390 | 0.14 | 0.18 | 0.44 | / | / | / | 0.22 | 0.29 | 0.45 | / | / | / |
| rs11209013 | 1 | 67190603 | 0.14 | 0.18 | 0.44 | <0.01 | 0.32 | 0.99 | -0.17 | 0.22 | 0.43 | 0.30 | 0.58 | 0.61 |
| rs11221335 | 11 | 128516011 | -0.14 | 0.20 | 0.47 | 0.61 | 0.36 | 0.09 | / | / | / | 1.08 | 0.64 | 0.09 |
| rs11236797 | 11 | 76588605 | <0.01 | 0.09 | 0.98 | -0.19 | 0.17 | 0.27 | -0.26 | 0.12 | 0.02 | <0.01 | 0.31 | 0.99 |
| rs112936798 | 1 | 39336709 | -0.17 | 0.28 | 0.54 | 0.36 | 0.34 | 0.29 | 0.38 | 0.38 | 0.33 | -0.38 | 0.59 | 0.52 |
| rs1131095 | 3 | 49676792 | -0.10 | 0.09 | 0.25 | -0.21 | 0.16 | 0.19 | 0.01 | 0.11 | 0.95 | -0.17 | 0.28 | 0.55 |
| rs11548656 | 16 | 81883307 | 0.27 | 0.16 | 0.09 | 0.20 | 0.25 | 0.42 | 0.17 | 0.24 | 0.49 | 0.06 | 0.46 | 0.90 |
| rs11581607 | 1 | 67242007 | -0.04 | 0.04 | 0.29 | -0.02 | 0.09 | 0.86 | 0.01 | 0.05 | 0.80 | -0.12 | 0.17 | 0.47 |
| rs11669299 | 19 | 10385945 | 0.11 | 0.16 | 0.49 | 0.37 | 0.30 | 0.21 | / | / | / | 0.96 | 0.54 | 0.07 |
| rs11677002 | 2 | 28391534 | -0.19 | 0.15 | 0.20 | 0.18 | 0.27 | 0.50 | -0.05 | 0.20 | 0.82 | -0.79 | 0.48 | 0.10 |
| rs11734570 | 4 | 38586832 | -0.17 | 0.21 | 0.41 | -0.74 | 0.36 | 0.04 | 0.06 | 0.33 | 0.85 | -0.27 | 0.65 | 0.68 |
| rs11768365 | 7 | 6505557 | 0.26 | 0.20 | 0.21 | 0.28 | 0.35 | 0.41 | 0.25 | 0.27 | 0.36 | -0.26 | 0.62 | 0.67 |
| rs117981694 | 12 | 40428296 | -0.16 | 0.13 | 0.23 | 0.06 | 0.22 | 0.79 | 0.60 | 0.16 | <0.01 | 0.54 | 0.41 | 0.19 |
| rs12136659 | 1 | 172875110 | -0.27 | 0.19 | 0.15 | -0.23 | 0.31 | 0.45 | 0.23 | 0.23 | 0.31 | -0.61 | 0.57 | 0.28 |
| rs1250573 | 10 | 79282718 | -0.29 | 0.15 | 0.06 | 0.14 | 0.27 | 0.59 | 0.08 | 0.25 | 0.75 | -1.03 | 0.48 | 0.03 |

| | | | | | | | | | | | | | | |
|---|---|---|---|---|---|---|---|---|---|---|---|---|---|---|
| rs1268339 | 1 | 1280044 | 0.07 | 0.21 | 0.73 | -0.23 | 0.30 | 0.45 | -0.22 | 0.28 | 0.44 | 0.27 | 0.53 | 0.61 |
| rs12825700 | 12 | 68099200 | -0.19 | 0.11 | 0.08 | -0.19 | 0.20 | 0.35 | 0.09 | 0.13 | 0.50 | 0.15 | 0.36 | 0.69 |
| rs12936409 | 17 | 39887396 | -0.01 | 0.10 | 0.90 | 0.31 | 0.18 | 0.08 | 0.14 | 0.12 | 0.23 | 0.29 | 0.32 | 0.37 |
| rs1297264 | 21 | 15443698 | 0.05 | 0.09 | 0.57 | -0.14 | 0.18 | 0.43 | 0.37 | 0.12 | <0.01 | -0.22 | 0.31 | 0.48 |
| rs1317209 | 1 | 19813543 | 0.12 | 0.15 | 0.44 | -0.24 | 0.26 | 0.36 | 0.02 | 0.25 | 0.94 | 0.39 | 0.46 | 0.40 |
| rs1336900 | 1 | 150706557 | 0.03 | 0.16 | 0.87 | -0.05 | 0.30 | 0.86 | 0.25 | 0.28 | 0.36 | -0.12 | 0.54 | 0.83 |
| rs13422838 | 2 | 186638119 | -0.35 | 0.20 | 0.07 | -0.99 | 0.43 | 0.02 | / | / | / | 0.97 | 0.76 | 0.21 |
| rs140933577 | 13 | 40262133 | <0.01 | 0.18 | 1.00 | / | / | / | / | / | / | / | / | / |
| rs143210366 | 6 | 31524576 | 0.10 | 0.13 | 0.45 | -0.19 | 0.19 | 0.33 | / | / | / | -0.23 | 0.37 | 0.53 |
| rs1445004 | 5 | 40414317 | -0.02 | 0.08 | 0.83 | -0.10 | 0.15 | 0.49 | 0.02 | 0.10 | 0.84 | 0.88 | 0.27 | <0.01 |
| rs145568234 | 6 | 32279268 | 0.06 | 0.12 | 0.60 | -0.36 | 0.25 | 0.15 | 0.10 | 0.10 | 0.31 | 0.07 | 0.48 | 0.89 |
| rs1456896 | 7 | 50264865 | 0.08 | 0.17 | 0.62 | 0.28 | 0.32 | 0.38 | -0.30 | 0.21 | 0.15 | 0.89 | 0.56 | 0.12 |
| rs149169037 | 7 | 20537675 | -0.09 | 0.19 | 0.63 | / | / | / | / | / | / | / | / | / |
| rs154873 | 20 | 59279977 | -0.04 | 0.18 | 0.81 | 0.20 | 0.32 | 0.52 | 0.04 | 0.26 | 0.87 | 0.01 | 0.57 | 0.98 |
| rs1558619 | 2 | 102315090 | -0.20 | 0.16 | 0.22 | 0.14 | 0.30 | 0.64 | 0.31 | 0.21 | 0.13 | 0.35 | 0.53 | 0.50 |
| rs16940202 | 16 | 85980635 | -0.19 | 0.16 | 0.22 | 0.49 | 0.32 | 0.12 | 0.01 | 0.19 | 0.97 | 0.54 | 0.56 | 0.34 |
| rs17656349 | 5 | 150226431 | 0.02 | 0.19 | 0.90 | -0.04 | 0.35 | 0.91 | 0.25 | 0.31 | 0.41 | -0.18 | 0.63 | 0.78 |
| rs1864239 | 15 | 79913012 | / | / | / | -0.01 | 0.04 | 0.86 | <0.01 | 0.14 | 0.98 | 0.06 | 0.07 | 0.42 |
| rs194746 | 14 | 68816170 | -0.13 | 0.16 | 0.41 | <0.01 | 0.30 | 1.00 | -0.28 | 0.21 | 0.18 | 0.67 | 0.54 | 0.21 |
| rs212402 | 6 | 159051263 | 0.43 | 0.20 | 0.03 | 0.50 | 0.37 | 0.18 | 0.17 | 0.27 | 0.51 | -1.19 | 0.66 | 0.07 |
| rs2301127 | 16 | 11273620 | -0.68 | 0.18 | <0.01 | 0.26 | 0.33 | 0.42 | 0.19 | 0.22 | 0.39 | 2.17 | 0.58 | <0.01 |
| rs2384352 | 16 | 11273620 | -0.08 | 0.15 | 0.61 | 0.22 | 0.28 | 0.42 | -0.25 | 0.13 | 0.05 | 1.26 | 0.49 | 0.01 |
| rs2413583 | 22 | 39263768 | -0.08 | 0.11 | 0.44 | -0.19 | 0.23 | 0.40 | -0.25 | 0.28 | 0.37 | 0.09 | 0.40 | 0.82 |
| rs243505 | 17 | 148738247 | -0.12 | 0.18 | 0.50 | -0.04 | 0.32 | 0.91 | / | / | / | 0.01 | 0.58 | 0.99 |
| rs2593855 | 3 | 71126344 | 0.03 | 0.18 | 0.85 | / | / | / | / | / | / | / | / | / |
| rs2836881 | 21 | 39094373 | -0.02 | 0.09 | 0.84 | -0.20 | 0.18 | 0.26 | -0.01 | 0.12 | 0.90 | -0.20 | 0.32 | 0.53 |
| rs28374519 | 16 | 28478021 | -0.03 | 0.14 | 0.85 | 0.05 | 0.24 | 0.83 | / | / | / | 0.38 | 0.43 | 0.38 |
| rs2838517 | 21 | 44193942 | 0.03 | 0.11 | 0.80 | / | / | / | 0.07 | 0.14 | 0.59 | / | / | / |

| | | | | | | | | | | | | | | |
|---|---|---|---|---|---|---|---|---|---|---|---|---|---|---|
| rs3024493 | 1 | 206770623 | 0.15 | 0.10 | 0.14 | 0.07 | 0.18 | 0.70 | -0.03 | 0.13 | 0.84 | 0.12 | 0.33 | 0.71 |
| rs341295 | 5 | 112513193 | -0.01 | 0.20 | 0.98 | -0.36 | 0.36 | 0.32 | 0.10 | 0.32 | 0.75 | 0.64 | 0.64 | 0.32 |
| rs34140409 | 6 | 31955877 | 0.13 | 0.15 | 0.39 | -0.09 | 0.33 | 0.78 | / | / | / | 1.64 | 0.61 | 0.01 |
| rs35171809 | 6 | 167019278 | -0.18 | 0.13 | 0.15 | 0.24 | 0.23 | 0.31 | -0.35 | 0.19 | 0.06 | -0.97 | 0.42 | 0.02 |
| rs3792111 | 2 | 233271044 | -0.01 | 0.10 | 0.89 | 0.14 | 0.18 | 0.44 | -0.02 | 0.12 | 0.88 | -0.01 | 0.32 | 0.98 |
| rs3820330 | 1 | 19815920 | 0.07 | 0.17 | 0.68 | -0.18 | 0.31 | 0.56 | 0.08 | 0.23 | 0.73 | -0.71 | 0.55 | 0.20 |
| rs3829110 | 9 | 136374746 | -0.08 | 0.09 | 0.39 | -0.19 | 0.16 | 0.25 | 0.20 | 0.11 | 0.06 | -0.01 | 0.29 | 0.97 |
| rs3850378 | 14 | 87951173 | 0.01 | 0.16 | 0.92 | / | / | / | -0.12 | 0.19 | 0.55 | / | / | / |
| rs3897234 | 13 | 26967893 | 0.28 | 0.17 | 0.10 | -0.08 | 0.30 | 0.79 | -0.14 | 0.22 | 0.51 | 0.27 | 0.54 | 0.61 |
| rs4256018 | 20 | 6113242 | -0.17 | 0.20 | 0.38 | -0.08 | 0.41 | 0.84 | 0.49 | 0.25 | 0.05 | -0.07 | 0.73 | 0.92 |
| rs4276914 | 1 | 155169753 | -0.11 | 0.17 | 0.53 | 0.12 | 0.32 | 0.70 | -0.27 | 0.22 | 0.22 | 0.60 | 0.57 | 0.30 |
| rs4380956 | 8 | 125516832 | -0.13 | 0.16 | 0.39 | 0.05 | 0.31 | 0.88 | -0.04 | 0.19 | 0.85 | 1.75 | 0.55 | <0.01 |
| rs4676408 | 2 | 240634984 | -0.08 | 0.14 | 0.57 | <0.01 | 0.26 | 0.98 | -0.07 | 0.18 | 0.72 | 0.44 | 0.46 | 0.34 |
| rs4807569 | 19 | 1123379 | 0.09 | 0.13 | 0.52 | -0.07 | 0.23 | 0.76 | 0.10 | 0.21 | 0.65 | -0.19 | 0.42 | 0.65 |
| rs4957256 | 5 | 40207040 | 0.02 | 0.15 | 0.88 | 0.35 | 0.24 | 0.15 | <0.01 | 0.20 | 0.99 | 0.52 | 0.42 | 0.22 |
| rs503734 | 3 | 101304904 | -0.12 | 0.20 | 0.55 | 0.26 | 0.37 | 0.48 | -0.20 | 0.26 | 0.44 | 1.08 | 0.65 | 0.10 |
| rs55946629 | 2 | 43624107 | <0.01 | 0.16 | 0.98 | 0.39 | 0.36 | 0.28 | -0.05 | 0.22 | 0.82 | 0.10 | 0.63 | 0.88 |
| rs56062135 | 15 | 67163292 | -0.14 | 0.12 | 0.25 | 0.26 | 0.21 | 0.20 | -0.06 | 0.14 | 0.70 | 0.22 | 0.37 | 0.55 |
| rs56116661 | 13 | 188683372 | -0.33 | 0.18 | 0.07 | -0.31 | 0.30 | 0.30 | -0.22 | 0.28 | 0.45 | 0.59 | 0.54 | 0.27 |
| rs56235845 | 5 | 177371039 | -0.22 | 0.17 | 0.19 | / | / | / | / | / | / | / | / | / |
| rs5754100 | 22 | 21561877 | -0.27 | 0.14 | 0.06 | 0.09 | 0.21 | 0.67 | / | / | / | -0.70 | 0.38 | 0.07 |
| rs5763793 | 22 | 30130643 | 0.04 | 0.19 | 0.84 | 0.16 | 0.35 | 0.66 | 0.15 | 0.24 | 0.53 | 0.25 | 0.62 | 0.69 |
| rs6017342 | 20 | 44436388 | 0.24 | 0.13 | 0.08 | 0.06 | 0.22 | 0.78 | -0.06 | 0.15 | 0.68 | -0.32 | 0.39 | 0.42 |
| rs6062496 | 20 | 63697746 | 0.08 | 0.11 | 0.48 | 0.30 | 0.19 | 0.11 | -0.31 | 0.13 | 0.02 | 0.31 | 0.34 | 0.37 |
| rs6063502 | 20 | 50339058 | 0.09 | 0.21 | 0.66 | -0.84 | 0.35 | 0.02 | 0.38 | 0.33 | 0.25 | -0.14 | 0.63 | 0.83 |
| rs62126610 | 19 | 33257277 | -0.19 | 0.14 | 0.17 | / | / | / | / | / | / | / | / | / |
| rs62183956 | 2 | 218181399 | 0.35 | 0.18 | 0.05 | -0.05 | 0.33 | 0.88 | -0.45 | 0.23 | 0.04 | 0.61 | 0.58 | 0.30 |
| rs62324212 | 4 | 122639784 | 0.27 | 0.16 | 0.09 | -0.32 | 0.29 | 0.27 | -0.04 | 0.22 | 0.86 | 0.99 | 0.52 | 0.05 |

| | | | | | | | | | | | | | | |
|---|---|---|---|---|---|---|---|---|---|---|---|---|---|---|
| rs62378712 | 5 | 159187037 | -0.20 | 0.20 | 0.32 | -0.26 | 0.40 | 0.51 | -0.16 | 0.28 | 0.57 | -0.25 | 0.70 | 0.72 |
| rs62408218 | 6 | 90222139 | -0.25 | 0.17 | 0.15 | -0.82 | 0.35 | 0.02 | 0.05 | 0.22 | 0.82 | -0.63 | 0.62 | 0.31 |
| rs62482552 | 7 | 100924735 | -0.24 | 0.20 | 0.22 | -0.32 | 0.35 | 0.36 | 0.23 | 0.27 | 0.39 | -0.83 | 0.63 | 0.18 |
| rs6457681 | 6 | 32805720 | 0.03 | 0.10 | 0.79 | / | / | / | 0.24 | 0.14 | 0.08 | / | / | / |
| rs6579807 | 5 | 150907283 | -0.09 | 0.16 | 0.55 | -0.05 | 0.30 | 0.86 | 0.17 | 0.20 | 0.38 | -1.24 | 0.53 | 0.02 |
| rs6584282 | 10 | 99526738 | 0.06 | 0.09 | 0.48 | -0.31 | 0.17 | 0.06 | 0.15 | 0.11 | 0.17 | -0.06 | 0.30 | 0.83 |
| rs6740847 | 2 | 181443625 | -0.03 | 0.15 | 0.82 | -0.22 | 0.28 | 0.43 | -0.04 | 0.21 | 0.84 | 0.82 | 0.50 | 0.10 |
| rs6873866 | 5 | 96912106 | -0.05 | 0.15 | 0.74 | / | / | / | / | / | / | / | / | / |
| rs6933404 | 6 | 137638098 | 0.45 | 0.20 | 0.02 | -0.13 | 0.38 | 0.73 | -0.34 | 0.24 | 0.16 | -0.19 | 0.66 | 0.77 |
| rs714910 | 17 | 34290246 | -0.02 | 0.16 | 0.92 | 0.06 | 0.28 | 0.82 | -0.27 | 0.20 | 0.18 | 0.41 | 0.50 | 0.41 |
| rs7190426 | 16 | 23844532 | -0.07 | 0.19 | 0.72 | -0.33 | 0.42 | 0.43 | 0.10 | 0.26 | 0.69 | 0.14 | 0.75 | 0.85 |
| rs7256518 | 19 | 10515699 | 0.03 | 0.19 | 0.87 | 0.14 | 0.36 | 0.70 | 0.08 | 0.23 | 0.72 | 1.15 | 0.64 | 0.07 |
| rs72852162 | 2 | 145486323 | / | / | / | 0.16 | 0.33 | 0.64 | / | / | / | 0.36 | 0.59 | 0.55 |
| rs744166 | 17 | 42362183 | -0.06 | 0.20 | 0.76 | -0.04 | 0.23 | 0.88 | 0.18 | 0.16 | 0.26 | -0.88 | 0.41 | 0.03 |
| rs749910 | 16 | 50724938 | 0.02 | 0.12 | 0.84 | -0.04 | 0.18 | 0.82 | 0.13 | 0.10 | 0.17 | 0.26 | 0.32 | 0.42 |
| rs7532133 | 1 | 160881744 | -0.03 | 0.19 | 0.86 | -0.08 | 0.34 | 0.81 | 0.07 | 0.23 | 0.76 | 0.02 | 0.61 | 0.97 |
| rs755374 | 5 | 159402286 | -0.03 | 0.09 | 0.69 | 0.13 | 0.16 | 0.41 | -0.03 | 0.11 | 0.78 | 0.04 | 0.28 | 0.89 |
| rs7608697 | 2 | 60977506 | -0.09 | 0.10 | 0.37 | 0.21 | 0.19 | 0.25 | -0.33 | 0.12 | 0.01 | 0.56 | 0.33 | 0.09 |
| rs76286777 | 2 | 24972708 | 0.07 | 0.17 | 0.69 | 0.76 | 0.37 | 0.04 | -0.07 | 0.21 | 0.73 | 0.55 | 0.65 | 0.40 |
| rs78771661 | 2 | 22129337 | -0.30 | 0.18 | 0.10 | 0.16 | 0.35 | 0.64 | -0.02 | 0.22 | 0.91 | 0.52 | 0.60 | 0.38 |
| rs7918084 | 10 | 92669710 | 0.27 | 0.19 | 0.16 | -0.33 | 0.36 | 0.35 | -0.12 | 0.25 | 0.62 | -1.35 | 0.63 | 0.03 |
| rs80262450 | 18 | 12818923 | -0.18 | 0.14 | 0.20 | 0.19 | 0.24 | 0.45 | -0.10 | 0.18 | 0.57 | -0.75 | 0.44 | 0.08 |
| rs938650 | 8 | 128540294 | -0.04 | 0.19 | 0.85 | 0.30 | 0.36 | 0.41 | -0.29 | 0.23 | 0.21 | 0.43 | 0.66 | 0.51 |
| rs9934775 | 16 | 50349166 | -0.03 | 0.16 | 0.86 | -0.26 | 0.33 | 0.42 | 0.10 | 0.27 | 0.72 | -0.84 | 0.57 | 0.14 |
| All - Inverse variance weighted | | | -0.03 | 0.01 | 0.03 | -0.01 | 0.02 | 0.58 | 0.01 | 0.02 | 0.50 | 0.08 | 0.05 | 0.10 |
| All - MR Egger | | | -0.02 | 0.03 | 0.66 | -0.01 | 0.04 | 0.76 | 0.05 | 0.04 | 0.24 | 0.03 | 0.09 | 0.75 |

AD: Alzheimer's disease; ALS: amyotrophic lateral sclerosis; Chr: chromosome; IBD: inflammatory bowel disease; IVW: Inverse variance weighted; MS: multiple sclerosis; PD: Parkinson's disease; SE: standard error; SNP: single-nucleotide polymorphism.

**Supplementary Table S3. Mendelian randomization (MR) analysis for the causality ulcerative colitis (UC) and Crohn's disease (CD) on neurodegenerative disorders.**

| | | | | Mendelian randomization | | | | Heterogeneity | |
|---|---|---|---|---|---|---|---|---|---|
| **Exposure** | **Outcome** | **Method** | **SNPs** | **OR** | **LL** | **UL** | ***P*** | **Q** | **Q_*P*** |
| CD | ALS | IVW | 78 | 0.97 | 0.95 | 1.00 | 0.02 | 77.55 | 0.46 |
| | | MR Egger | 78 | 1.02 | 0.96 | 1.09 | 0.52 | 75.01 | 0.51 |
| | | Weighted median | 78 | 0.99 | 0.96 | 1.02 | 0.50 | | |
| | | Weighted mode | 78 | 1.00 | 0.94 | 1.05 | 0.87 | | |
| | | MR PERSSO | 78 | 0.97 | 0.95 | 0.99 | 0.02 | | |
| | AD | IVW | 74 | 1.00 | 0.96 | 1.05 | 0.94 | 49.58 | 0.45 |
| | | MR Egger | 74 | 1.02 | 0.90 | 1.15 | 0.76 | 45.98 | 0.56 |
| | | Weighted median | 74 | 1.02 | 0.96 | 1.08 | 0.56 | | |
| | | Weighted mode | 74 | 1.03 | 0.93 | 1.13 | 0.59 | | |
| | PD | IVW | 71 | 1.01 | 0.98 | 1.05 | 0.43 | 94.55 | 0.03 |
| | | MR Egger | 71 | 1.09 | 1.00 | 1.18 | 0.05 | 89.96 | 0.05 |
| | | Weighted median | 71 | 1.01 | 0.96 | 1.06 | 0.67 | | |
| | | Weighted mode | 71 | 1.00 | 0.94 | 1.07 | 0.97 | | |
| | MS | IVW | 74 | 1.03 | 0.93 | 1.15 | 0.53 | 148.87 | <0.01 |
| | | MR Egger | 74 | 0.98 | 0.74 | 1.30 | 0.91 | 148.58 | <0.01 |
| | | Weighted median | 74 | 1.07 | 0.94 | 1.21 | 0.30 | | |
| | | Weighted mode | 74 | 1.07 | 0.90 | 1.28 | 0.46 | | |
| UC | ALS | IVW | 52 | 1.01 | 0.98 | 1.03 | 0.68 | 40.34 | 0.86 |
| | | MR Egger | 52 | 1.02 | 0.94 | 1.11 | 0.60 | 40.18 | 0.84 |
| | | Weighted median | 52 | 1.00 | 0.96 | 1.04 | 0.94 | | |
| | | Weighted mode | 52 | 1.02 | 0.95 | 1.10 | 0.55 | | |
| | AD | IVW | 50 | 0.99 | 0.94 | 1.04 | 0.73 | 49.58 | 0.45 |
| | | MR Egger | 50 | 1.14 | 0.98 | 1.33 | 0.10 | 45.98 | 0.56 |
| | | Weighted median | 50 | 0.95 | 0.88 | 1.04 | 0.26 | | |
| | | Weighted mode | 50 | 0.90 | 0.72 | 1.11 | 0.32 | | |
| | PD | IVW | 49 | 0.99 | 0.96 | 1.03 | 0.78 | 51.30 | 0.35 |
| | | MR Egger | 49 | 1.01 | 0.91 | 1.13 | 0.80 | 51.15 | 0.31 |
| | | Weighted median | 49 | 1.01 | 0.96 | 1.06 | 0.78 | | |
| | | Weighted mode | 49 | 1.02 | 0.94 | 1.10 | 0.68 | | |
| | MS | IVW | 49 | 1.11 | 0.95 | 1.30 | 0.20 | 121.31 | <0.01 |
| | | MR Egger | 49 | 0.93 | 0.54 | 1.60 | 0.78 | 120.14 | <0.01 |
| | | Weighted median | 49 | 1.03 | 0.89 | 1.19 | 0.67 | | |
| | | Weighted mode | 49 | 0.92 | 0.71 | 1.18 | 0.49 | | |

AD: Alzheimer's disease; ALS: amyotrophic lateral sclerosis; CD: Crohn's disease; IVW: Inverse variance weighted; LL: lower limits of odds ratio; OR: Odds ratio; PD: Parkinson's disease; SNP: single-nucleotide polymorphism; UC: ulcerative colitis; MS: multiple sclerosis; UL: upper limits of odds ratio.

**Supplementary Table S4. Assessment of pleiotropy.**

| | | MR-Egger intercept | | | MR PRESSO global test | |
|---|---|---|---|---|---|---|
| **Exposure** | **Outcome** | **Intercept** | **SE** | ***P*** | **RSSobs** | ***P*** |
| IBD | ALS | -0.00201 | 0.004186 | 0.632572 | 115.3298 | 0.211 |
| | AD | -0.00011 | 0.005126 | 0.983122 | 91.2978 | 0.600 |
| | PD | -0.00558 | 0.005688 | 0.329295 | 116.9115 | 0.033 |
| | MS | 0.00958 | 0.012046 | 0.428585 | 161.6244 | <0.001 |
| UC | ALS | -0.00252 | 0.006287 | 0.690204 | 42.1793 | 0.865 |
| | AD | -0.02302 | 0.012122 | 0.063533 | 53.1721 | 0.383 |
| | PD | -0.00328 | 0.008716 | 0.708321 | 53.6505 | 0.306 |
| | MS | 0.027634 | 0.040836 | 0.501911 | 126.469 | <0.001 |
| CD | ALS | -0.00814 | 0.005105 | 0.114797 | 79.8534 | 0.461 |
| | AD | -0.02302 | 0.012122 | 0.063533 | 53.1721 | 0.402 |
| | PD | -0.01352 | 0.007206 | 0.064890 | 97.5203 | 0.033 |
| | MS | 0.00877 | 0.023651 | 0.712005 | 152.7905 | <0.001 |

AD: Alzheimer's disease; ALS: amyotrophic lateral sclerosis; Chr: chromosome; IBD: inflammatory bowel disease; IVW: Inverse variance weighted; MS: multiple sclerosis; PD: Parkinson's disease; SE: standard error.

**Supplementary Table S5. Mendelian randomization (MR) analysis for the causality of neurodegenerative disorders on inflammatory bowel disease (IBD), including ulcerative colitis (UC) and Crohn's disease (CD).**

| Exposure | Outcome | Method | SNPs | Mendelian randomization | | | |
|---|---|---|---|---|---|---|---|
| | | | | OR | LL | UL | *P* |
| ALS | IBD | IVW | 4 | 1.00 | 0.89 | 1.12 | 1.00 |
| | | MR Egger | 4 | 0.92 | 0.70 | 1.20 | 0.59 |
| | | Weighted median | 4 | 1.03 | 0.90 | 1.19 | 0.65 |
| | | Weighted mode | 4 | 1.05 | 0.91 | 1.20 | 0.57 |
| | UC | IVW | 4 | 0.98 | 0.82 | 1.17 | 0.78 |
| | | MR Egger | 4 | 0.77 | 0.54 | 1.10 | 0.29 |
| | | Weighted median | 4 | 1.04 | 0.87 | 1.23 | 0.69 |
| | | Weighted mode | 4 | 1.04 | 0.85 | 1.27 | 0.75 |
| | CD | IVW | 4 | 1.01 | 0.86 | 1.18 | 0.94 |
| | | MR Egger | 4 | 1.04 | 0.67 | 1.62 | 0.87 |
| | | Weighted median | 4 | 1.06 | 0.90 | 1.25 | 0.50 |
| | | Weighted mode | 4 | 1.08 | 0.90 | 1.29 | 0.47 |
| AD | IBD | IVW | 2 | 1.01 | 0.98 | 1.04 | 0.58 |
| | | MR Egger | / | / | / | / | / |
| | | Weighted median | / | / | / | / | / |
| | | Weighted mode | / | / | / | / | / |
| | UC | IVW | 2 | 1.00 | 0.96 | 1.03 | 0.82 |
| | | MR Egger | / | / | / | / | / |
| | | Weighted median | / | / | / | / | / |
| | | Weighted mode | / | / | / | / | / |
| | CD | IVW | 2 | 1.00 | 0.97 | 1.04 | 0.78 |
| | | MR Egger | / | / | / | / | / |
| | | Weighted median | / | / | / | / | / |
| | | Weighted mode | / | / | / | / | / |
| PD | IBD | IVW | 17 | 1.03 | 0.99 | 1.08 | 0.19 |
| | | MR Egger | 17 | 1.07 | 0.96 | 1.19 | 0.25 |
| | | Weighted median | 17 | 1.03 | 0.97 | 1.10 | 0.32 |
| | | Weighted mode | 17 | 1.03 | 0.94 | 1.12 | 0.56 |
| | UC | IVW | 17 | 1.03 | 0.97 | 1.09 | 0.30 |
| | | MR Egger | 17 | 1.02 | 0.89 | 1.18 | 0.74 |
| | | Weighted median | 17 | 1.06 | 0.98 | 1.14 | 0.15 |
| | | Weighted mode | 17 | 1.09 | 0.97 | 1.22 | 0.16 |
| | CD | IVW | 17 | 1.04 | 0.98 | 1.10 | 0.24 |
| | | MR Egger | 17 | 1.08 | 0.94 | 1.24 | 0.31 |
| | | Weighted median | 17 | 1.03 | 0.95 | 1.12 | 0.48 |
| | | Weighted mode | 17 | 1.01 | 0.88 | 1.15 | 0.89 |
| MS | IBD | IVW | 4 | 1.11 | 1.04 | 1.18 | <0.01 |
| | | MR Egger | 4 | 1.54 | 1.02 | 2.33 | 0.18 |
| | | Weighted median | 4 | 1.11 | 1.07 | 1.14 | <0.01 |

| | | | | | | |
|---|---|---|---|---|---|---|
| | Weighted mode | 4 | 1.14 | 1.10 | 1.18 | 0.01 |
| UC | IVW | 4 | 1.18 | 1.03 | 1.34 | 0.02 |
| | MR Egger | 4 | 2.71 | 1.19 | 6.18 | 0.14 |
| | Weighted median | 4 | 1.11 | 1.04 | 1.18 | <0.01 |
| | Weighted mode | 4 | 1.26 | 1.20 | 1.33 | <0.01 |
| CD | IVW | 4 | 1.04 | 0.98 | 1.11 | 0.23 |
| | MR Egger | 4 | 0.86 | 0.48 | 1.54 | 0.66 |
| | Weighted median | 4 | 1.02 | 0.98 | 1.07 | 0.28 |
| | Weighted mode | 4 | 1.02 | 0.98 | 1.06 | 0.41 |

IBD: inflammatory bowel disease; AD: Alzheimer’s disease; ALS: amyotrophic lateral sclerosis; CD: Crohn’s disease; IVW: Inverse variance weighted; LL: lower limits of odds ratio; MS: multiple sclerosis; OR: Odds ratio; PD: Parkinson’s disease; SNP: single-nucleotide polymorphism; UC: ulcerative colitis; UL: upper limits of odds ratio.

**Supplementary Table S6. Genetic correlation between inflammatory bowel disease (IBD) and amyotrophic lateral sclerosis (ALS).**

| ρ | SE | *P* | Correlation |
|---|---|---|---|
| -0.0472 | 0.0057 | $8.60\times10^{-17}$ | -0.3255 |

**Supplementary Table S7. Functional enrichment of shared risk loci.**

| Category | Pathway (ID) | *P* value | Adjusted *P* value |
|---|---|---|---|
| Biological Process | regulation of ER to Golgi vesicle-mediated transport (GO:0060628) | 0.0013 | 0.0149 |
| | negative regulation of autophagosome assembly (GO:1902902) | 0.0019 | 0.0149 |
| | regulation of establishment of protein localization (GO:0070201) | 0.0022 | 0.0149 |
| | negative regulation of macroautophagy (GO:0016242) | 0.0038 | 0.0149 |
| | regulation of protein transport (GO:0051223) | 0.0041 | 0.0149 |
| | regulation of autophagosome assembly (GO:2000785) | 0.0052 | 0.0149 |
| | negative regulation of organelle assembly (GO:1902116) | 0.0052 | 0.0149 |
| | regulation of transport (GO:0051049) | 0.0055 | 0.0149 |
| | regulation of intracellular transport (GO:0032386) | 0.0058 | 0.0149 |
| | COPII vesicle coating (GO:0048208) | 0.0094 | 0.0180 |
| | vesicle coating (GO:0006901) | 0.0094 | 0.0180 |
| | vesicle targeting, rough ER to cis-Golgi (GO:0048207) | 0.0094 | 0.0180 |
| | COPII-coated vesicle budding (GO:0090114) | 0.0104 | 0.0185 |
| | retrograde vesicle-mediated transport, Golgi to endoplasmic reticulum (GO:0006890) | 0.0126 | 0.0191 |
| | retrograde transport, endosome to Golgi (GO:0042147) | 0.0131 | 0.0191 |
| | post-Golgi vesicle-mediated transport (GO:0006892) | 0.0132 | 0.0191 |
| | regulation of vesicle-mediated transport (GO:0060627) | 0.0143 | 0.0193 |
| | cytosolic transport (GO:0016482) | 0.0173 | 0.0221 |
| | endoplasmic reticulum to Golgi vesicle-mediated transport (GO:0006888) | 0.0274 | 0.0332 |
| | protein-containing complex assembly (GO:0065003) | 0.0395 | 0.0454 |
| Cellular Component | Golgi-associated vesicle (GO:0005798) | 0.0070 | 0.0298 |
| | cis-Golgi network (GO:0005801) | 0.0085 | 0.0298 |
| Molecular Function | syntaxin binding (GO:0019905) | 0.0092 | 0.0092 |

GO, Gene Ontology.
